# Onset of Alzheimer disease in apolipoprotein ɛ4 carriers is earlier in butyrylcholinesterase K variant carriers

**DOI:** 10.1101/2024.01.02.24300718

**Authors:** Roger M. Lane, Taher Darreh-Shori, Candice Junge, Dan Li, Qingqing Yang, Amanda L. Edwards, Danielle L. Graham, Katrina Moore, Catherine J. Mummery

## Abstract

**Background:** We wished to examine the impact of the K-variant of *butyrylcholinesterase* (*BCHE-K*) carrier status on age-at-diagnosis of Alzheimer disease (AD) in *APOE4* carriers.

**Methods:** In 45 patients, aged 50-74 years, with cerebrospinal fluid (CSF) biomarker confirmed mild AD, recruited into a clinical trial (NCT03186989), baseline demographics, disease characteristics, and biomarkers were evaluated by *BCHE-K* and *APOE4* allelic status.

**Results:** In *APOE4* carriers (N = 33), mean age-at-diagnosis of AD in *BCHE-K* carriers (n = 11) was 6.4 years earlier than in *BCHE-K* noncarriers (n = 22, *P <* .001, ANOVA). In *APOE4* noncarriers (N = 12) there was no similar influence of *BCHE-K*. In *APOE4* carriers with versus those without *BCHE-K*, mean age-at-baseline was over 6 years earlier and accompanied by slightly higher amyloid and tau accumulations. A predominant amyloid, limited tau pathophysiology, and limbic-amnestic phenotype was exemplified by *APOE4* homozygotes with *BCHE-K*. Multiple regression analyses demonstrated association of amyloid accumulation with *APOE4* carrier status (*P <* .029), larger total brain ventricle volume (*P <* .021), less synaptic injury (Ng, *P <* .001), and less tau (p-tau_181_, *P <* .005). In contrast, tau pathophysiology was associated with more neuroaxonal damage (NfL, *P* = .002), more synaptic injury (Ng, *P <* .001), and higher levels of glial activation (YKL-40, *P* = .01).

**Conclusion:** Findings concern the genetic architecture of prognosis in early AD, that is fundamental for patients and the design of clinical trials, and that is less well established than the genetics of susceptibility. In mild AD patients aged less than 75 years, the mean age-at-diagnosis of AD in *APOE4* carriers was reduced by over 6 years in *BCHE-K* carriers versus noncarriers. Functional activation of glia may explain much of the effects of *APOE4* and *BCHE-K* on the phenotype of early AD.

## Background

The cholinergic hypothesis of AD states that selective loss of cholinergic neurons, arising from basal forebrain nuclei, and decreased levels of the neurotransmitter, acetylcholine (ACh), trigger neurodegeneration and cognitive impairment (1). Corticolimbic cholinergic denervation may be evident at early stages of AD (2, 3). The failure of this circuitry is inextricably linked with cognitive deficits in memory, learning, attention, and processing speed (4). Synaptic release of ACh initiates cholinergic neurotransmission and is rapidly terminated by acetylcholinesterase (AChE) located in the synapse. The availability of ACh in cholinergic synapses is deficient in AD and can be increased with acetylcholinesterase inhibitors (AChE-Is) (5). The cholinergic system produces both rapid focal synaptic signaling and slow diffuse extracellular signaling though alpha 7 nicotinic ACh receptors (α7-nAChRs), and both act to control glial cell reactivity and functional state (6). Glial cells provide for the homeostasis and neuroprotection of the central nervous system (CNS), and if this functionality is deficient, amyloid pathology can accumulate (7). In contrast, tau tangle pathology is more strongly correlated with glial activation, and microglial and astrocyte activation may better predict the spatiotemporal spread of tau tangles (8, 9).

*Apolipoprotein E* (*APOE*) encodes ApoE, the major intercellular lipid carrier in the CNS that is mainly produced by astrocytes, reactive microglia, vascular mural cells, and choroid plexus cells (10, 11). The *APOE4* polymorphism is the major genetic risk factor for sporadic AD (12). In *APOE4* carriers, functional glial responses to clear Aβ are deficient and favor the accumulation of amyloid pathology (7). Cerebral amyloid accumulation begins earlier in life in *APOE4* carriers than in noncarriers (13, 14).

Butyrylcholinesterase (BuChE), along with AChE, is involved in the enzymatic breakdown of both synaptic and extracellular ACh (15, 16). Astrocytes secrete BuChE and the ACh synthesizing enzyme, choline acetyltransferase (ChAT), to maintain a steady state equilibrium of hydrolysis and synthesis of extracellular ACh (17). In addition to particular populations of neurons in the amygdala and hippocampus (18), BuChE is localized in glia, myelin, and endothelial cells, and continues to increase in concentration with age, especially in the deep cortex and white matter (19). Aβ, ApoE, and BuChE interact with each other to influence the catalytic activity of BuChE, and these entities are also prominent constituents of amyloid plaque (20, 21). For example, CSF ApoE protein, in a concentration- and polymorphism-dependent manner, profoundly alters the catalytic functioning and stability of CSF BuChE in patients with mild AD; an interaction that is Aβ concentration-dependent (20, 22). *BCHE* genotype and CSF BuChE activity are correlated with markers of glial activation in early AD (23, 24). Lower BuChE activity is associated with higher amyloid accumulation in patients with mild AD (24).

The most common single-nucleotide polymorphism (SNP) of *BCHE*, the Kalow-variant (*BCHE-K;* 3q26.1-3q26.2; nucleotide G1615A, codon A539T; rs1803274), is carried by 18-35% of individuals in Western populations (25–28). In *APOE4* carriers, reduced BuChE activity is more marked in *BCHE-K* carriers, with a *BCHE-K* allele dose-dependent reduction in BuChE activity and lowering of glial activation markers (24, 29). The *BCHE-K* and *APOE4* alleles significantly interact to reduce the age-at-onset of AD (30), and to increase the likelihood of progression from mild cognitive impairment (MCI) to AD (27, 31) and from cognitively unimpaired older individuals to early AD (26). Carriers of both *APOE4* and *BCHE-K* alleles in the MCI stage of AD have a limbic-amnestic phenotype and progress most rapidly in the mild stage of AD, where they are the only genotype group with a significant response to AChE-I treatment (27, 31–33).

The primary objective of this cross-sectional analysis of mild AD patients aged less than 75 years was to evaluate *BCHE-K* effects on age-at-onset of AD in *APOE4* carriers. This analysis found evidence for an amyloid-accumulating phenotype in carriers of both *APOE4* and *BCHE-K*, apparently dependent on levels of glial activation, that mediated an earlier age-at-onset of AD. These findings link amyloid, innate immune, and cholinergic mechanisms in the underlying disease process.

## Methods

This trial (NCT03186989) was conducted in accordance with Good Clinical Practice Guidelines of the International Council for Harmonisation, and according to the ethical principles outlined in the Declaration of Helsinki. The complete study (34) was approved by relevant ethics committees. Written informed consent was provided by the participants.

### Study eligibility criteria

Eligible study participants were between the ages of 50 and 74; had probable mild AD (amnestic or non-amnestic), defined by a Mini-Mental State Examination score (MMSE) (35) of 20-27, inclusive (representative of mild AD), and either a Clinical Dementia Rating (36) Overall Global Score of 1, or a Global Score of 0.5 with a Memory Score of 1; a CSF pattern of low Aβ_1-42_ (≤ 1200 pg/ml), elevated total-tau (> 200 pg/ml) and p-tau (> 18 pg/ml), and a total-tau to Aβ_1-42_ ratio > 0.28; and a diagnosis of probable AD based on National Institute of Aging-Alzheimer Association (NIA-AA) criteria (37).

### Assessments

CSF from participants was analyzed for markers of amyloid accumulation (inversely indexed by Aβ_42_), tau pathophysiology (p-tau_181_), neuroaxonal degeneration (NfL), synaptic injury (Ng), glial activation (YKL-40). The CSF analytes and assays are detailed in Table 1. Of 46 patients entering the study, 45 were characterized for common SNPs of *APOE* (ie, *APOE4*, *APOE3*, and *APOE2*), and *BCHE* rs1803274 (ie, *BCHE-K*).

**Table 1.** CSF biomarkers of AD pathology, glial activation, inflammation, neuroaxonal damage, and synaptic injury.

| CSF analyte | Biology indexed |
| --- | --- |
| **Aβ<sub>42</sub>** (β-amyloid 1-42)<br>Elecsys β-amyloid 1-42 CSF performed at Roche Diagnostics, Indianapolis, IN | Parenchymal amyloid accumulation |
| **p-tau<sub>181</sub>** (tau phosphorylated at threonine 181)<br>Elecsys Phospho-Tau (181P) CSF performed at Roche Diagnostics, Indianapolis, IN | Tau tangle pathology |
| **NfL\*** (neurofilament light chain)<br>Uman, performed at Immunologix, Tampa, FL | Neuroaxonal degeneration |
| **Ng\*** (neurogranin)<br>Euroimmune, performed at Immunologix, Tampa, FL) | Postsynaptic injury |
| **YKL-40\*** (chitinase-3-like protein 1)<br>ELLA (Protein Simple), performed at Immunologix, Tampa, FL) | Microglial and astrocyte activation marker (39) |

\*All CSF samples for NfL, Ng, and YKL-40 were tested using a single batch of reagents.

The study required 3-dimensional (3D) T1-weighted structural magnetic resonance imaging (MRI) scans of the head, volumetric analyses calculated using VivoQuant^TM^, which is comprised of a preprocessing module and a multi-atlas segmentation module, followed by visual inspection and manual editing if needed (38). The mean baseline ventricular volume and hippocampal volume were expressed as a percentage of the total intracranial volume (%TIV).

### Statistical analysis

Patient baseline characteristics were summarized according to genotype and sex (Tables 2-4). Quantitative assessments were summarized using descriptive statistics, including number of patients, mean, and standard deviation. Qualitative assessments were summarized using frequency counts and percentages. The exact test was used to examine the Hardy-Weinberg equilibrium (HWE) in the distribution of *APOE* and *BCHE* alleles in the study population. All exact tests were performed using the R package “Hardy Weinberg” (40). The HWE *P* value, which measures the strength of evidence against the null hypothesis that the distribution does not follow Hardy-Weinberg equilibrium, was provided. A large *P* value is consistent with the distribution following HWE.

**Table 2.** Mild AD phenotype across genotype groups defined by *APOE4* allele frequency and for *APOE4* homozygotes by the presence of *BCHE-K* alleles.

|  | Mean value for variable (SD) | 2 *APOE4* &<br>*BCHE-K*<br>(N = 4) | 2 *APOE4*<br>(N = 10) | 1 *APOE4*<br>(N = 23) | No *APOE4*<br>(N = 12) | 1 or 2<br>*APOE4*<br>(N = 33) |
| --- | --- | --- | --- | --- | --- | --- |
| **Sex** | Female | 75% | 60% | 52% | 33% | 55% |
| **Onset of AD** | Age-at-diagnosis (yrs) | 60.8<br>(4.4) | 64.2<br>(5.4) | 65.2<br>(5.8) | 63.3<br>(7.4) | 64.9<br>(5.6) |
|  | Time since AD diagnosis at baseline (yrs) | 1.2<br>(1.2) | 1.2*<br>(1.2) | 1.4†<br>(1.3) | 1.3†<br>(1.5) | 1.4<br>(1.2) |
| **Neuroimaging** | Hippocampal vol., % of ICV | 0.26<br>(0.05) | 0.24*<br>(0.04) | 0.25*<br>(0.04) | 0.28†<br>(0.03) | 0.25<br>(0.04) |
|  | Ventricular vol., % of ICV | 1.8<br>(0.73) | 1.8†<br>(0.53) | 3.3*<br>(1.32) | 2.7*<br>(1.08) | 2.8<br>(1.31) |
| **CSF markers** | Aβ<sub>42</sub>, pg/mL | 554<br>(111) | 581*<br>(110) | 700†<br>(188) | 799†<br>(180) | 664<br>(175) |
|  | p-tau<sub>181</sub>, pg/mL | 34.0<br>(11.8) | 35.2†<br>(9.3) | 41.1*<br>(15.2) | 41.3*<br>(13.8) | 39.3<br>(13.8) |
|  | NfL, pg/mL | 1003<br>(265) | 1039†<br>(319) | 1383*<br>(438) | 1399*<br>(296) | 1279<br>(432) |
|  | Ng, pg/mL | 493<br>(188) | 482†<br>(120) | 546*<br>(271) | 526*<br>(235) | 527<br>(235) |
|  | YKL-40, ng/mL | 190<br>(86.0) | 202†<br>(63.8) | 267*<br>(114.9) | 318*<br>(161.9) | 247<br>(105.6) |
| **Cognition** | MMSE Total (0-30) | 24.3<br>(2.5) | 24.4†<br>(2.1) | 23.3*<br>(2.3) | 23.7*<br>(2.2) | 23.6<br>(2.3) |
|  | MMSE Memory (0-6) | 4.0<br>(1.4) | 4.1*<br>(1.0) | 4.0*<br>(1.1) | 4.6†<br>(1.2) | 4.1<br>(1.1) |
|  | MMSE Visual Construction (0-1) | 0.8<br>(0.5) | 0.8†<br>(0.4) | 0.7†<br>(0.5) | 0.5*<br>(0.5) | 0.7<br>(0.5) |
|  | RBANS Total (40-160) | 72.8<br>(8.3) | 73.4†<br>(10.5) | 64.0*<br>(11.2) | 68.5*<br>(12.3) | 66.8<br>(11.7) |
|  | RBANS Delayed Memory (40-154) | 56.0<br>(19.3) | 52.8*<br>(12.5) | 48.0*<br>(7.4) | 66.3†<br>(22.5) | 49.5<br>(9.3) |
|  | RBANS Visuospatial/Constructional (40-154) | 101.8<br>(28.8) | 100.8†<br>(20.0) | 81.1*<br>(19.8) | 79.9*<br>(21.7) | 87.1<br>(21.6) |

Across only the *APOE4* allele-frequency groups: \* Numerically more pathology or more clinical impairment; † Numerically less pathology or less clinical impairment.
ICV, intracranial volume; MMSE, Mini-Mental Status Examination; RBANS, Repeatable Battery for the Assessment of Neuropsychological Status; SD, standard deviation.

Both analysis of variance (ANOVA) and analysis of covariance (ANCOVA) were used to test whether the mean age-at-diagnosis of AD (primary analysis) or the mean age-at-baseline differed across two or more genotype groups, ie, by *BCHE-K* carrier status in *APOE4* carriers, and in *APOE4* homozygotes and heterozygotes. When the ANCOVA model was applied, the model included *BCHE-K* carrier status and sex as factors, and baseline MMSE total score as covariate. Prior to performing ANOVA and ANCOVA, the normality assumption of residuals was tested using the Kolmogorov-Smirnov test. If significant departures from normality were observed, the Wilcoxon Rank Sum test was applied. Both ANOVA and ANCOVA were applied to test baseline CSF Aβ_42_ across two or more genotype groups. When ANCOVA was applied, age-at-baseline was included as an additional covariate in the model. If the normality assumption was not satisfied, both ANOVA and ANCOVA model were fitted to the log-transformed data. Box plots were used to visualize data by group.

Relationships between CSF Aβ_42_, CSF p-tau_181_, other CSF biomarkers, and brain volumes (in terms of total hippocampal and total brain ventricular volumes as %TIV) were explored in the overall population and in each genotype group in a simple linear correlation analysis with a Pearson correlation coefficient. The squared Pearson correlation coefficient (*R*^2^) and *P* value were provided in the correlation analysis and interpreted by descriptors to indicate the strength of the relationship. Correlation coefficients with a magnitude of 0.81 < *R*^2^ < 1 indicate variables were very highly/strongly correlated; 0.49 < *R*^2^ < 0.81 indicates highly/strongly correlated; 0.25 < *R*^2^ < 0.49 indicates moderately correlated; 0.09 < *R*^2^ < 0.25 indicates a low/weak correlation; and R^2^ < 0.09 indicates negligible correlation. Scatterplots with a simple linear regression line were produced to depict the relationships between two quantitative variables.

Multiple regression analysis was also utilized to assess the functional relationships between the biomarker of interest and amyloid and tau pathophysiology. A multiple regression model was applied with CSF Aβ_42_ and CSF p-tau_181_ as the response variables, and *APOE4*, *BCHE-K*, age-at-baseline, sex, baseline MMSE total score, and the biomarker of interest (ie, CSF p-tau_181_ or Aβ_42_, NfL, Ng, YKL-40; total hippocampal volume, total brain ventricular volume) as independent variables. This determined the strength of association of CSF Aβ_42_ or CSF p-tau with parameters of interest, in conjunction with the other independent variables included in the model.

## Results

The study was conducted at 12 centers in Canada, Finland, Germany, the Netherlands, Sweden, and the UK between August 2017 and February 2020. 102 patients were assessed for eligibility. Of these, 56 were excluded, and one enrolled participant did not provide genetic test results. The study sample comprised 45 participants with a mean age of 65.8 years and a mean baseline MMSE total score of 23.6.

The *APOE4* allele was carried by 73% (33) of participants; 22% (10) homozygotes and 51% (23) heterozygotes (Table 2). The *BCHE-K* allele was carried by 36% (16) of participants; 7% (3) homozygotes and 29% (13) heterozygotes (Table S1). Of the 16 *BCHE-K* carriers, 69% (11) were *APOE4* carriers (Table 3). All non-*APOE4* alleles were *APOE3*, except for one participant with an *APOE2/3* genotype, who also carried one *BCHE-K* allele. Both the distribution of *APOE* genotypes (HWE exact *P* value = 1) and *BCHE* genotypes (HWE exact *P* value = .376) were consistent with Hardy-Weinberg equilibrium.

**Table 3.** Mild AD phenotype across genotype groups defined by *APOE4* and *BCHE-K* carrier status.

|  | Mean value for variable (SD) | *E4 & K*<br>(N = 11) | *E4 & No-K*<br>(N = 22) | *No-E4 & K*<br>(N = 5) | *No-E4 & No-K*<br>(N = 7) |
| --- | --- | --- | --- | --- | --- |
| **Sex** | Female | 64% | 50% | 40% | 29% |
| **Onset of AD** | Age-at-AD diagnosis (yrs) | 60.6*<br>(6.1) | 67.0†<br>(4.0) | 63.9<br>(7.6) | 62.8<br>(7.8) |
|  | Age-at-baseline (yrs) | 62.1*<br>(5.9) | 68.3†<br>(4.0) | 64.8<br>(7.3) | 64.4<br>(6.6) |
|  | Time since AD diagnosis at baseline (yrs) | 1.5†<br>(1.3) | 1.3*<br>(1.2) | 0.9*<br>(0.7) | 1.6†<br>(1.9) |
| **Neuroimaging** | Hippocampal vol., % of ICV | 0.26*<br>(0.05) | 0.24*<br>(0.03) | 0.28†<br>(0.02) | 0.28†<br>(0.03) |
|  | Ventricular vol., % of ICV | 2.9*<br>(1.54) | 2.8†<br>(1.23) | 2.3†<br>(0.97) | 2.9*<br>(1.15) |
| **CSF markers** | Aβ<sub>42</sub>, pg/mL | 641*<br>(139) | 676*<br>(193) | 755†<br>(195) | 831†<br>(176) |
|  | p-tau<sub>181</sub>, pg/mL | 40.4*<br>(12.2) | 38.7†<br>(14.8) | 42.5*<br>(15.8) | 40.4*<br>(13.5) |
|  | NfL, pg/mL | 1392*<br>(498) | 1222†<br>(395) | 1316†<br>(160) | 1458*<br>(366) |
|  | Ng, pg/mL | 568*<br>(188) | 506†<br>(257) | 540*<br>(245) | 515†<br>(247) |
|  | YKL-40, ng/mL | 247†<br>(143.3) | 247†<br>(85.0) | 351*<br>(164.5) | 293*<br>(168.5) |
| **Cognition** | MMSE Total (0-30) | 23.8†<br>(2.5) | 23.5*<br>(2.3) | 23.4*<br>(2.4) | 23.9†<br>(2.3) |
|  | MMSE Memory (0-6) | 3.9*<br>(1.0) | 4.1*<br>(1.1) | 4.2*<br>(1.3) | 4.9†<br>(1.2) |
|  | MMSE Visual Construction (0-1) | 0.7†<br>(0.5) | 0.7†<br>(0.5) | 0.8†<br>(0.4) | 0.3*<br>(0.5) |
|  | RBANS Total (40-160) | 65.5*<br>(10.5) | 67.5†<br>(12.4) | 72.0†<br>(13.4) | 66.0*<br>(11.8) |
|  | RBANS Delayed Memory (40-154) | 52.7*<br>(13.7) | 47.8*<br>(5.9) | 69.0†<br>(26.2) | 64.3†<br>(21.5) |
|  | RBANS Visuospatial/Constructional (40-154) | 90.4†<br>(25.6) | 85.4†<br>(19.8) | 79.4*<br>(21.1) | 80.3*<br>(23.9) |

\* Numerically more pathology or more clinical impairment; † Numerically less pathology or less clinical impairment; ICV, intracranial volume; MMSE, Mini-Mental Status Examination; Repeatable Battery for the Assessment of Neuropsychological Status; SD, standard deviation.

### Age-at-diagnosis of AD in *APOE4* carriers was reduced in carriers of *BCHE-K* alleles

There were no significant *APOE4* carrier or allele frequency-associated differences on the participants’ age-at-diagnosis of AD or on their age-at-baseline of study (Table 2). *APOE4* homozygotes (n = 10) had a mean age-at-diagnosis of AD of 64.2 years versus heterozygotes (n = 23) of 65.2 years, versus noncarriers (n = 12) of 63.3 years. In contrast, *BCHE-K* homozygotes (n = 3) had a lower mean age-at-diagnosis of AD of 59.4 years versus heterozygotes (n = 13) of 62.2 years, versus noncarriers (n = 29) of 66.0 years (*P* = .048, ANOVA) (Table S1).

In the primary analysis of this investigation, amongst *APOE4* carriers, *BCHE-K* carriers (n=11) versus noncarriers (n=22) were associated with differences in mean age-at-diagnosis of AD of 6.4 years (60.6 versus 67.0; *P <* .001, ANOVA; *P* = .001, ANCOVA) (Figure 1A; Table 3), and mean age-at-baseline of study of 6.2 years (62.1 versus 68.3; *P* = .001, ANOVA; *P* = .002, ANCOVA) (Figure 1C). The difference in mean age-at-diagnosis of AD in *BCHE-K* carriers versus noncarriers was similar in *APOE4* heterozygotes at 6.7 years (60.5 versus 67.3 years) and homozygotes at 5.7 years (60.8 versus 66.4 years) (*P* = .013, ANOVA; *P* = .019, ANCOVA) (Figure 1B). Significant differences of a similar magnitude were also seen in age-at-baseline of study (Figure 1D).

**Figure 1.**
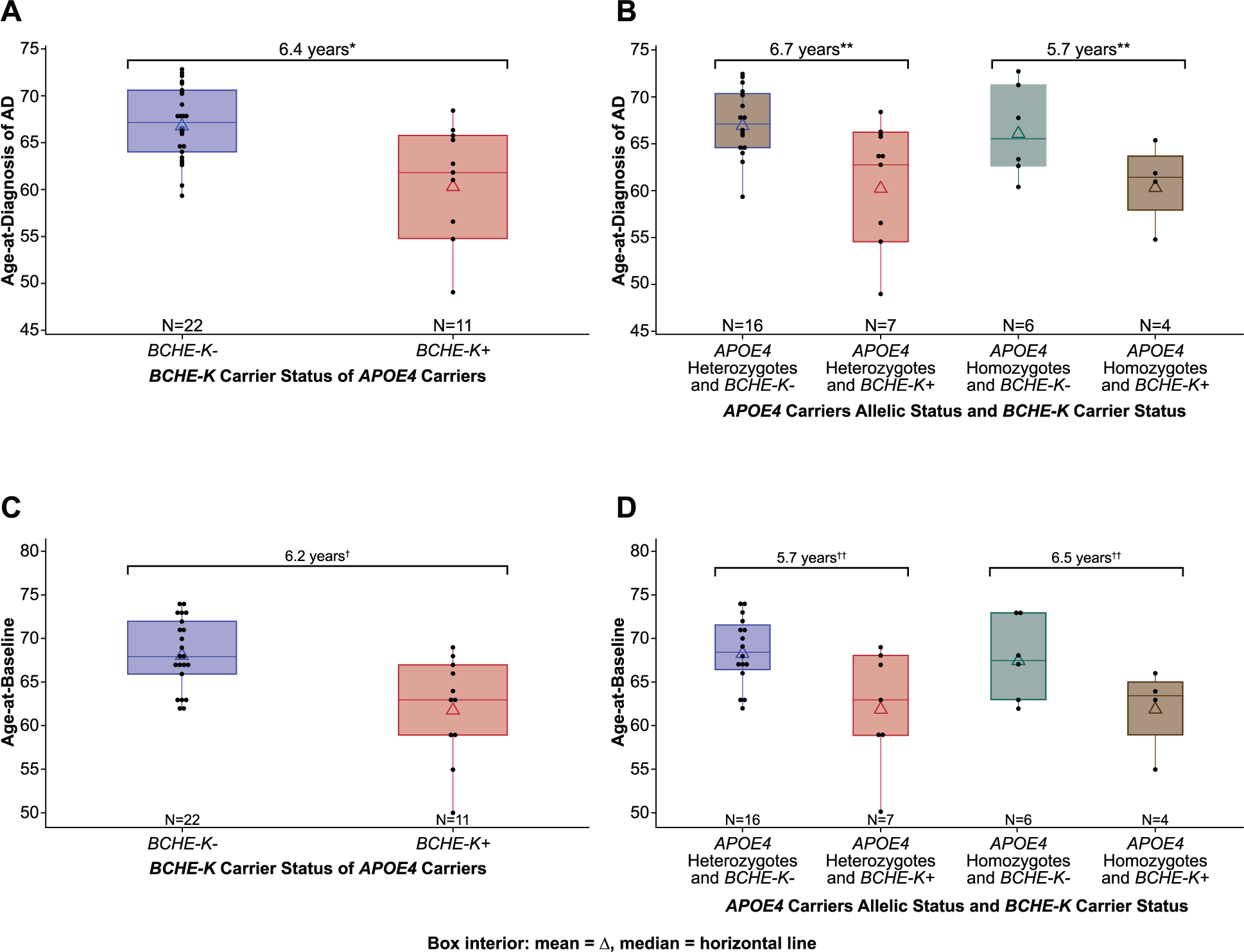
Age-at-diagnosis of AD and age-at-baseline by *BCHE-K* carrier status in *APOE4* carriers, homozygotes, and heterozygotes. A. Age-at-diagnosis of AD in *APOE4* carriers by *BCHE-K* carrier status B. Age-at-diagnosis of AD in *APOE4* homozygotes and heterozygotes by *BCHE-K* carrier status C. Age-at-baseline of study in *APOE4* carriers by *BCHE-K* carrier status D. Age-at-baseline of study in *APOE4* homozygotes and heterozygotes by *BCHE-K* carrier status * *P <* .001, ANOVA; *P* = .001, ANCOVA with *BCHE-K* carrier status and sex as factors and baseline MMSE total score as covariate for mean age-at-diagnosis of AD. ** *P* = .013, ANOVA; *P* = .019, ANCOVA with *APOE4* homozygotes and heterozygotes by *BCHE-K* carrier status and sex as factors and baseline MMSE total score as covariate for mean age-at-diagnosis of AD. † *P* = .001, ANOVA; *P* = .002, ANCOVA with *BCHE-K* carrier status and sex as factors and baseline MMSE total score as covariate for mean age-at-study-baseline. †† *P* = .015, ANOVA; *P* = .025, ANCOVA with *APOE4* homozygotes and heterozygotes by *BCHE-K* carrier status and sex as factors and baseline MMSE total score as covariate for mean age-at-study-baseline.

In *APOE4* noncarriers (N = 12), the mean age-at-diagnosis and age-at-baseline of the study were similar between *BCHE-K* carriers (n = 5) at 63.9 and 64.8 years, respectively, and noncarriers (n = 7) at 62.8 and 64.4 years, respectively (Table 3).

### In *APOE4* carriers with *BCHE-K*, younger mean age-at-baseline was accompanied by slightly higher amyloid and tau accumulations (Table 3)

Across genotype groups defined by *APOE4* and *BCHE-K* carrier status the proportion of female participants fell as the burden of *APOE4* and *BCHE-K* alleles diminished (Table 3). Carriers of both alleles had the earliest age-at-diagnosis, generally greater memory deficits, and the most amyloid accumulation. *APOE4* carriers with *BCHE-K* had slightly higher amyloid and tau accumulations than *APOE4* carriers without *BCHE-K*, despite a mean age-at-baseline that was 6.2 years earlier. In *APOE4* carriers, neurodegeneration and glial activation were similar between carriers and noncarriers of *BCHE-K* (Table 3). On the other hand, noncarriers of both *APOE4* and *BCHE-K* had the least amyloid pathology and multidomain cognitive deficits, with relatively less memory impairment. Individuals in these contrasting groups, defined by the presence or absence of both *APOE4* and *BCHE-K* alleles, had similar levels of tau and neurodegenerative pathology (Table 3).

### Higher *APOE4* allele frequency associates with amyloid pathology and the limbic-amnestic phenotype (Table 2)

Female participants exhibited a higher *APOE4* allele frequency than male participants. In *APOE4* carriers, there was a more temporo-limbic (hippocampal atrophy > ventricular expansion) and amnestic (memory > visuospatial impairment) phenotype relative to *APOE4* noncarriers (Table 2). *APOE4* carriers relative to noncarriers had higher levels of amyloid pathology (inversely indexed by the CSF Aβ_42_ = 664 versus 799 pg/mL, respectively; *P* = .028 ANOVA, *P* = .028 ANCOVA) (Table 2). *APOE4* homozygotes had the highest levels of amyloid pathology (CSF Aβ_42_ = 581 pg/mL) and the lowest levels of tau pathophysiology (CSF p-tau_181_ = 35.2 pg/mL), neuroaxonal damage (NfL), and synaptic injury (Ng). Glial activation (YKL-40) showed apparent *APOE4* allele frequency-dependent decreases to the lowest levels in homozygotes. Amyloid pathology was further increased in *APOE4* homozygotes with *BCHE-K* (CSF Aβ_42_ = 554 pg/mL), with further reductions of tau (CSF p-tau_181_ = 34.0 pg/mL), neurodegenerative pathology, and glial activation (Table 2).

### More amyloid pathology associated with less tau pathophysiology especially in carriers of both *APOE4* and *BCHE-K* (Figure 2)

Multiple regression analyses indicated that associations with amyloid pathology (inversely indexed by CSF Aβ_42_) included *APOE4* carrier status (*P <* .029), larger total brain ventricle volume (*P <* .021), less synaptic injury (Ng, *P <* .001), less tau (p-tau_181_, *P* = .005), and showed a trend for an association with less glial activation (*P* = .097). In simple linear correlation analyses in the overall population, amyloid pathology showed weak to moderate inverse correlations with tau pathophysiology, synaptic injury, and glial activation (Figures 2, 3B, and 3C). In carriers of both *APOE4* and *BCHE-K,* inverse correlations of amyloid with both synaptic injury and tau pathophysiology strengthened to moderate and weak inverse correlation with glial activation persisted but was not statistically significant (Figures 2, 3B, and 3C). In *APOE4* carriers without *BCHE-K*, inverse correlations of amyloid with tau pathophysiology were weak, and with synaptic injury and glial activation were moderate (Figures 2 and 3C).

**Figure 2.**
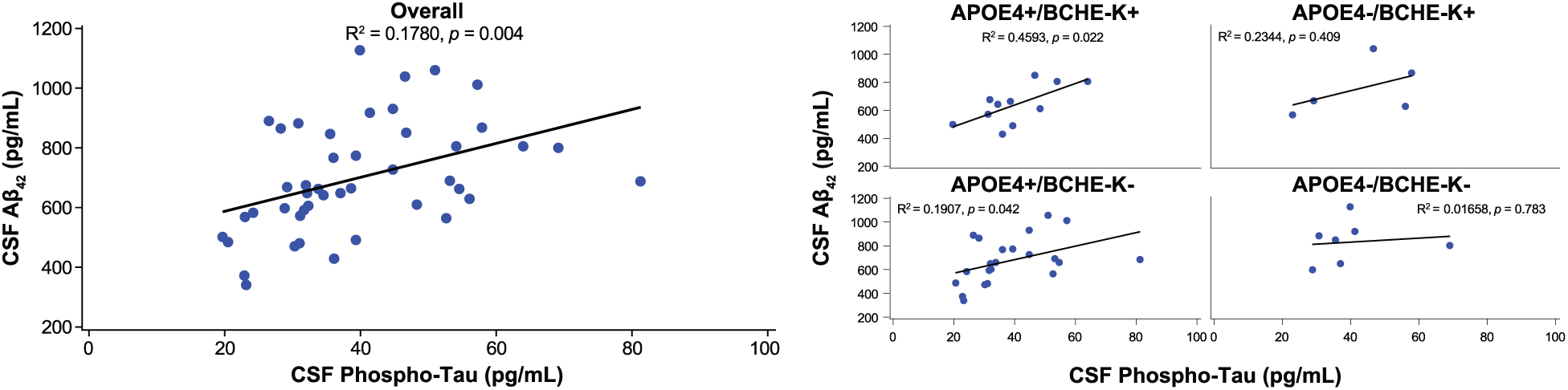
Correlations of CSF Aβ_42_ versus CSF p-tau_181_ in the overall population and in *APOE4* and *BCHE-K* subgroups. Simple linear correlation analyses: *R* square and *P* value were obtained by fitting a simple linear regression model.

**Figure 3.**
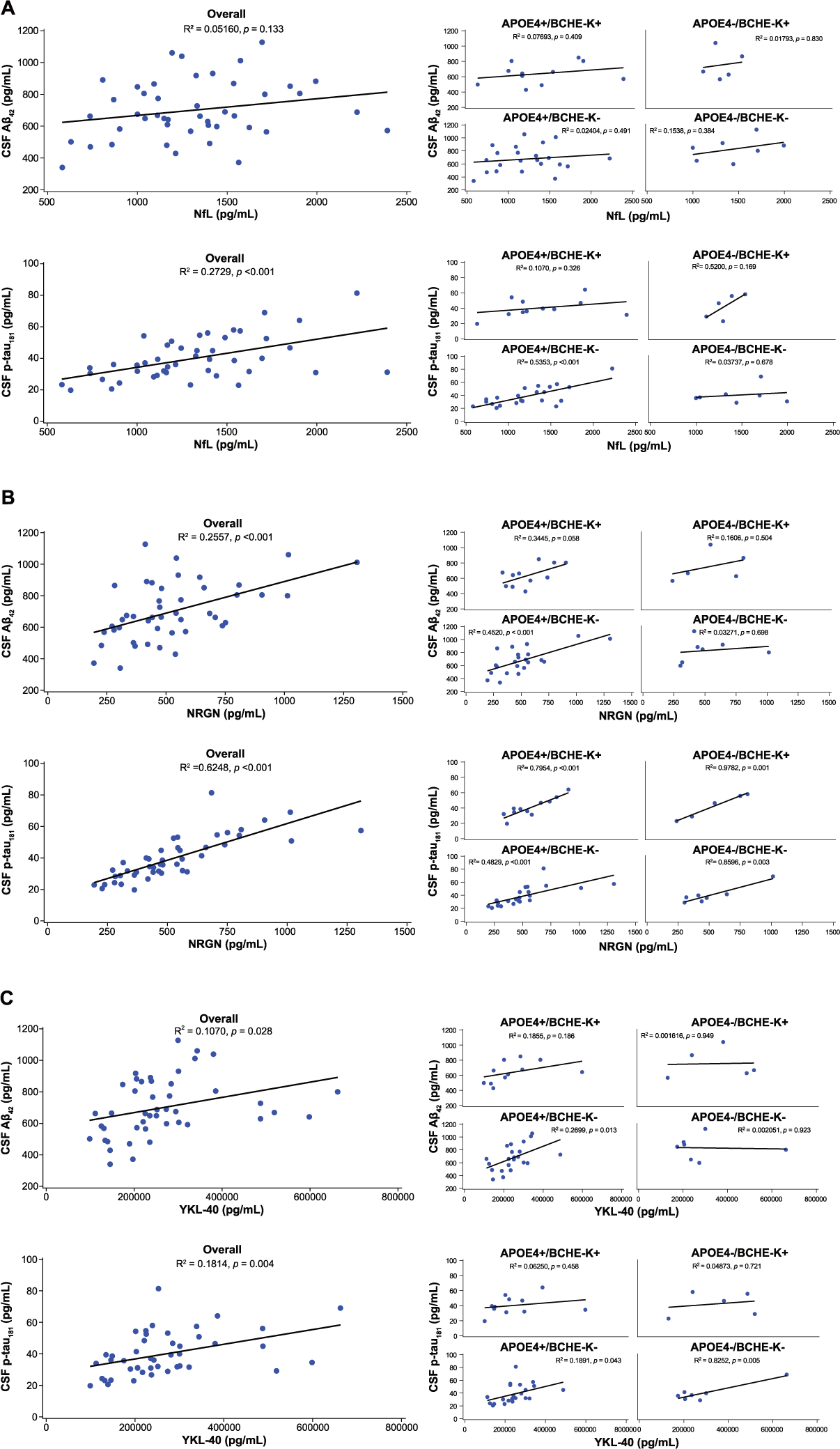
Correlations of neurodegenerative and glial activation markers with CSF Aβ_42_ and CSF p-tau_181_ in the overall study population and in genotype groups. A. CSF neurofilament light chain (NfL) (higher levels index more neuroaxonal injury) B. CSF neurogranin (Ng) (higher levels index more synaptic injury) C. CSF YKL-40 (higher levels index more glial activation) Simple linear correlation analyses: *R* square and *P* value were obtained by fitting a simple linear regression model. Only those analytes associated with either CSF Aβ_42_ or CSF p-tau_181_ in multiple regression analyses are shown.

### Tau pathophysiology associated with synaptic injury and glial activation, especially in noncarriers of both *APOE4* and *BCHE-K,* and with neuroaxonal degeneration, especially in *APOE4* carriers without *BCHE-K* (Figure 3)

Multiple regression analyses indicated associations with tau pathophysiology (p-tau_181_) with higher CSF levels of NfL (*P* = .002), Ng (*P <* .001), and YKL-40 (*P* = .01). Thus, more tau pathophysiology associated with neuroaxonal and synaptic degeneration, and glial activation. In simple linear regressions in the overall population, tau pathophysiology was moderately correlated with neuroaxonal damage, highly correlated with synaptic injury, and weakly correlated with glial activation (Figures 3A-C). Simple linear regressions showed correlations of tau pathophysiology with synaptic injury that was moderate in *APOE4* carriers without *BCHE-K*, strong in carriers of both *APOE4* and *BCHE-K*, and very strong in *APOE4* non-carrier subgroups (Figure 3B). Simple linear regression showed tau pathophysiology had a moderate correlation with the neuroaxonal degeneration in the overall group, and this strengthened to highly correlated in *APOE4* carriers without *BCHE-K* (Figure 3A). In simple linear regressions, correlation of glial activation with tau pathophysiology was weak in the overall population, weak in *APOE4* carriers without *BCHE-K*, and very strong in noncarriers of both *APOE4* and *BCHE-K* (Figure 3C). YKL-40 is proposed as a marker of tau pathology (41).

## Discussion

In this sample of clinically and pathologically characterized mild AD patients aged less than 75 years, *APOE4* carriers with *BCHE-K* had a mean age-at-diagnosis of AD 6.4 years earlier than in *APOE4* carriers without *BCHE-K* (Figure 1A). In *APOE4* carriers, slightly higher accumulations of amyloid and tau pathophysiology were present in *BCHE-K* carriers over 6 years earlier than in *BCHE-K* noncarriers (Table 3). The magnitude of this modifier effect may be due to a population age range in which the effects of *APOE4* on the AD phenotype are maximal and most modifiable. *APOE4* allele frequency-dependent effects on the risk-of-onset of AD are highest in the seventh decade, wane over 70 years of age, and are particularly reduced after 80 years of age (42, 43). The restricted age range for entry into the current study may also have been at least partly responsible for the lack of any significant *APOE4* allele frequency-dependent effects on age-at-diagnosis of AD or age-at-baseline (Table 2).

### Amyloid pathology may accumulate at an earlier age in *APOE4* carriers with *BCHE-K* alleles

*APOE4/E4* individuals start accumulating amyloid at an earlier age, followed by *APOE3/E4*, *APOE2/E4*, *APOE3/E3*, and *APOE2/E3* (13). The age of symptom onset may strongly correlate with the age at which an individual reaches a tipping point in fibrillar Aβ accumulation (44). In the current study, amyloid accumulation reached higher levels in genotype groups with lower levels of glial activation and an inverse correlation was observed between amyloid pathology and glial activation (Table 2; Figure 3C). Levels of amyloid pathology across genotype subgroups are compatible with *APOE4* allele frequency-dependent accumulation beginning at an earlier age with the earliest start in *APOE4* carriers with *BCHE-K* alleles, particularly in *APOE4* homozygotes with *BCHE-K* (Table 2).

### An age- and genotype-dependent phenotypic extreme of limbic-amnestic and amyloid-predominant mild AD is associated with low levels of glial activation (Figures 3 and 4; Table 2)

Optimally, glial activation may balance needs to attenuate amyloid accumulation and limit the spread of tau pathology (45, 46). This is an important determinant of the pathology mix and clinical features of early AD, including the age-of-onset of AD, and it links amyloid and tau pathology, cholinergic signaling, and glial activation (33).

**Figure 4.**
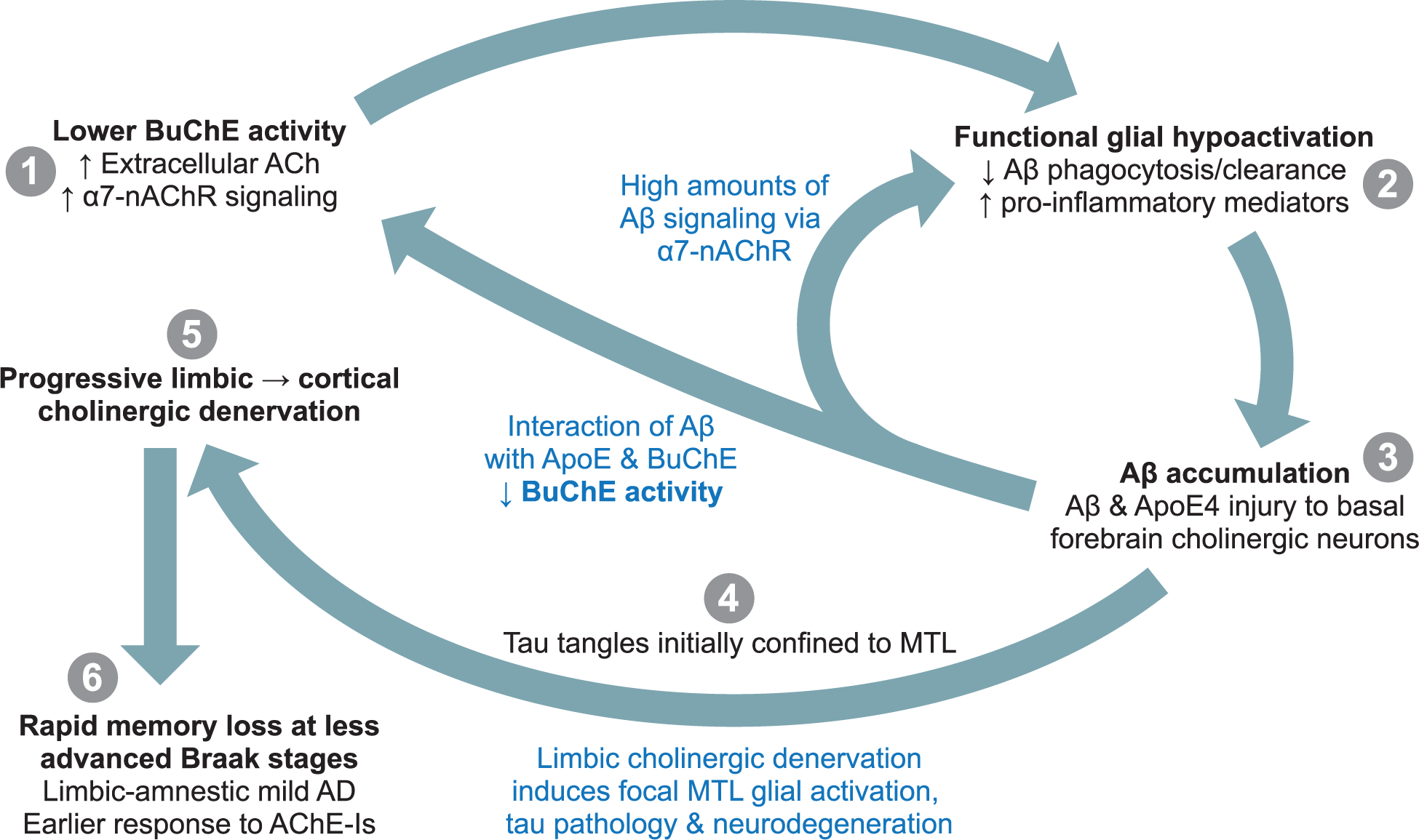
Amyloid pathology facilitating phenotype of mild AD exemplified by *APOE4* homozygotes with *BCHE-K* aged < 75 years. In preclinical and prodromal AD, lower BuChE activity, especially in *APOE4* homozygote *BCHE-K* carriers below 75 years of age, results in higher extracellular ACh and increased signaling through nAChR on glial cells (1). “*Functionally underactive*” glia with decreased phagocytic function and homeostatic responsiveness (2), impair Aβ clearance and induce earlier and greater amyloid accumulation (3). In prodromal and mild AD, limbic cholinergic denervation due to ApoE4-, Aβ- and tau-mediated damage to basal forebrain cholinergic neurons, removes the cholinergic “brake” on glia to increase glial activation, tau pathology and neurodegeneration in the MTL (4), but the spread of this pathology outside of the MTL is initially limited until cholinergic denervation has progressed to include other neocortical areas (5). Thus, high levels of amyloid accumulation develop at a younger age, and basal forebrain cholinergic denervation of MTL structures results in hippocampal atrophy and a rapidly progressing limbic-amnestic presentation in mild AD with good response to AChE-Is (6). The progression of corticolimbic cholinergic denervation to neocortical areas beyond the MTL results in the spread of glial activation, tau pathology, and neurodegeneration.

At one end of the age- and genotype- and disease stage-dependent spectrum of early AD is the limbic-amnestic and amyloid accumulation-predominant phenotype, exemplified *APOE4* homozygotes with *BCHE-K* alleles aged less than 75 years (Table 2). In *APOE4* carriers with *BCHE-K*, greater accumulation of Aβ pathology at younger ages is likely due to greater deficits in glial-mediated clearance mechanisms (7, 47) (Figures 3C and 4). These functionally underactive glia may produce proinflammatory cytokines and be classified as “inflammatory,” but their endolysosomal and phagocytic functions may be greatly reduced (48). Putatively lower BuChE activity in *BCHE-K* carriers with *APOE4* alleles results in higher extracellular ACh, further reduces the phagocytic function and responsiveness of glial cells, and further impairs Aβ clearance. While excessive cholinergic signaling encourages high levels of amyloid accumulation, the spread and accumulation of tau pathology is kept in check (Figures 2 and 4). However, in early AD, *APOE4* is associated with rapidly increasing tau pathology as the combination of ApoE4 and Aβ pathology induces sufficient degeneration of basal forebrain corticolimbic cholinergic neurons to release the cholinergic “brake” on glial activation in the medial temporal lobe (MTL) and eventually also in other cortical regions (Figure 4). Cholinergic denervation and tau pathology accumulating in the MTL results in a rapidly progressing limbic-amnestic presentation with response to AChE-I that is apparent in the mild stage of AD (32, 33). The focal MTL/limbic denervation is indexed by a more limbic-amnestic presentation (memory > visuospatial impairment, and hippocampal atrophy > ventricular expansion) (Tables 2 and 3).

### Both Aβ and ApoE modulate the cholinergic system

The cholinergic system acts to control glial reactivity and function though α7-nAChRs via rapid focal synaptic signaling and slow diffuse extracellular signaling (6). ApoE forms soluble and highly stable complexes with cholinesterase enzymes and Aβ, that can oscillate between slow and ultrafast ACh hydrolysis, depending on Aβ availability (29). Reduced cholinesterase activity and lowering of glial activation markers is seen in *APOE4* carriers, particularly in individuals with polymorphic variants of genes encoding cholinesterase enzymes with lower activity, such as *BCHE-K* (24, 29). In addition, lower cholesterol delivery by ApoE4 to the long and extensively arborized axons of the metabolically taxed cholinergic neurons requires them to expend energy on cholesterol synthesis and to consume acetyl coenzyme A (acetyl-CoA) (49). Acetyl-CoA is an essential substrate for the synthesis of both ACh and lipids essential for myelin formation and maintenance of cellular membranes (50). The ascending white matter projections of the basal forebrain cholinergic system may be particularly vulnerable to the combination of Aβ pathology and ApoE4 (51–53).

In a concentration- and aggregation-dependent manner, Aβ targets cholinergic synapses, signals through α7-nAChRs, and also influences the extracellular fluid equilibrium between breakdown and synthesis of ACh via effects on ACh-hydrolyzing capacity of cholinesterase (22) and choline acetyltransferase (ChAT) activity (54). In both aging and AD, the intraneuronal accumulation of oligomeric assemblies of Aβ_42_ is a relatively selective trait of basal forebrain cholinergic neurons (55, 56), and endocytic internalization of Aβ-nAChR complexes may underlie intracellular accumulation of Aβ_42_ and its neurotoxic consequences, such as tau phosphorylation (57). In AD, α7-nAChR expression on astrocytes is positively correlated with neuritic plaque burden (58). In an amyloid mouse model, loss of α7-nAChRs reduced Aβ_42_ plaque load, but increased levels of soluble Aβ_42_ oligomers and accelerated memory decline (59). Thus, Aβ and ApoE have physiological and disease roles in the tuning of cholinergic activity and in the vulnerability of the basal forebrain cholinergic system to degeneration, and these actions influence the functional status of cholinoceptive neuronal and nonexcitable cells (17).

### Hypofunctional glial-mediated clearance of Aβ likely underlies the “amyloid accumulating” phenotype exemplified by carriers of *APOE4* and *BCHE-K* aged < 75 years

A balanced activation of glia may be needed to stimulate Aβ clearance and avoid “hypofunctional” glia promoting amyloid accumulation versus “hyperactivated” glia that facilitate the dissemination of tau (45, 46, 60). In amyloid mouse models, inhibition of reactive astrogliosis increases Aβ_42_ plaque burden (61), whereas shifting microglia to an interferon-responsive state boosts ApoE expression, phagocytosis, containment of plaques, and lessens damage to nearby neurons and synapses (62). However, further shifting of microglia to an overactivated state may increase synaptic engulfment and accelerate the dissemination of tau pathology (63). Mouse models indicate that ApoE controls glial activation, with ApoE4 locking microglia in a homeostatic state with failure to clear neurodegenerative pathology (7, 47). Glia in *APOE4* mice demonstrate a significant decrease in phagocytic capacity (64). Microglial and astrocyte coverage of plaques is likely protective for surrounding neurons, and ApoE4 is associated with decreased coverage and more neuronal dystrophy (65–67).

*BCHE-K* carriers, who have lower levels of glial activation markers and higher levels of proinflammatory cytokines (24, 68), may exhibit deficient glial responses to neurodegeneration. Rapid and appropriately tuned changes in the catalytic activity of BuChE, and necessary adaptive changes in cholinoceptive nonexcitable cells may be more difficult to achieve with BuChE-K, particularly in *APOE4* carriers. Carriers of *APOE4* and *BCHE-K* may have more senescent microglia that are associated with blocked endolysosomal processing, impaired phagocytosis, and with acceleration of Aβ_42_ pathology (69). YKL-40 is a context-dependent modulator of glial phagocytic activity in both mice and humans (70). In the current study, *APOE4* noncarriers had higher mean levels of CSF YKL-40, at 318 ng/mL, relative to *APOE4* carriers at 247 ng/mL (Table 2). Mean levels were further reduced in *APOE4* homozygotes at 202 ng/mL and in *APOE4* homozygotes with *BCHE-K* alleles at 190 ng/mL (Table 2). This likely explains the inverse correlation between amyloid pathology and YKL-40 and synaptic injury seen in simple linear regression analyses overall and more specifically in *APOE4* carriers (Figure 3B and 3C). In addition, a weak correlation between YKL-40 and synaptic injury overall was evident in *BCHE-K* non-carriers (Figure S1). The correlation persisted as weak in *APOE4* carriers without *BCHE-K* and strengthened to high in non-carriers of both *APOE4* and *BCHE-K*.

Increases in functional glial activation with age might explain why *APOE4*-associated risk for AD reduces over the age range of 70-80 years, and why progression to dementia in prodromal AD carriers of both *APOE4* and *BCHE-K* is at least 2-fold higher in those aged below 75 years relative to older patients (33). In a longitudinal study of prodromal AD, progression to AD over 3-4 years in *APOE4* and *BCHE-K* carriers was 39% in participants aged less than 75 years and decreased to 18% in those aged 75 years or more (33). This was contrary to findings in the overall study, where progression to AD was greater in older participants (29%), relative to those aged less than 75 years (13%). Younger *APOE4* carriers have accelerated progression of hippocampal atrophy in prodromal and early stage AD, but in individuals more advanced in age or in progression of disease, any influence of *APOE4* on hippocampal atrophy is lost (71). Moreover, global cerebral atrophy in AD patients with a mean age of 70 years was reduced in an *APOE4* allele frequency-dependent manner (72), whereas in older patients, with a mean age of 80 years, atrophy was no different by genotype (71, 73).

### The “amyloid accumulating and initially tau spread limiting” phenotype supports the amyloid cascade hypothesis

Carriers of *APOE4* and *BCHE-K* with prodromal AD, in addition to progression rates to AD that inversely correlated with age, had the greatest hippocampal atrophy and declines in short- and long-term retrieval from verbal memory and overall cognitive impairment (27, 33). Likewise, in the current study of mild AD patients, *APOE4* carriers relative to noncarriers had greater memory deficits, hippocampal atrophy, and amyloid accumulation, and these findings in *APOE4* carriers occurred over 6 years earlier in *BCHE-K* carriers compared to noncarriers (Figure 1C, Table 3). The amyloid cascade hypothesis of AD implies that reaching the threshold for parenchymal amyloid positivity at an earlier age should drive secondary effector tau pathology, with an earlier age-at-onset of AD (74). Support for this hypothesis comes from slower progression of tau tangle accumulation and cognitive decline following antibody-induced removal of amyloid plaque below key threshold levels in patients with early symptomatic AD (75, 76). However, in the current study, correlations of amyloid pathology with tau pathophysiology in *APOE4* carriers were negative, particularly in carriers of both *APOE4* and *BCHE-K* (Figure 2). Across groups defined by *APOE4* allele frequency, *APOE4* homozygotes had the highest amyloid pathology and lowest tau and neurodegenerative pathology (Table 2). Amyloid pathology was further increased in *APOE4* homozygotes with *BCHE-K*, with further reduction of tau and neurodegenerative pathology (Table 2).

The inverse correlations of amyloid pathology with tau pathophysiology and synaptic injury may be a consequence of tau and neurodegenerative pathology localized to the MTL (which includes the entorhinal cortex, amygdala, and hippocampus). This MTL tau and neurodegenerative pathology may be responsible for transitioning *APOE4* and *BCHE-K* carriers into AD at an earlier age with less global tau and neurodegenerative pathology (Table 3; Figure 4) (77, 78). Such neuroanatomical distinctions are not discernible on CSF indices that summarize pathology changes across the brain. Spatial resolution requires tau tangle-ligand positron-emission tomography (tau-PET) neuroimaging. In the presence of global amyloid pathology in *APOE4* carriers, tau-PET indicates that tau pathology is more severe with a focal MTL distribution (78, 79). Furthermore, younger age is associated with PET-tau signal in the MTL in early AD *APOE4* carriers but not in *APOE4* noncarriers (78). Across the aging and AD spectrum, *APOE4* carriers present increased microglial activation relative to noncarriers in early Braak stage regions within the MTL and this microglial activation mediates Aβ-independent effects of *APOE4* on tau accumulation that are further associated with neurodegeneration and clinical impairment (80).

Degeneration of basal forebrain cholinergic neurons, that project to the MTL and other cortical structures, precedes and predicts longitudinal entorhinal/MTL degeneration (81, 82) and preclinical *APOE4* carriers exhibit the greatest loss of basal forebrain volume (83). The ascending neuronal projections of the basal forebrain cholinergic system may be particularly vulnerable to the combination of ApoE4-mediated glial hypofunction, high levels of Aβ, and tau pathologies (51–53). The impact of focal basal forebrain pathology is magnified as it causes widespread presynaptic cholinergic corticolimbic denervation. Both amyloid and tau pathologies may be required for substantial impairment of cholinergic synaptic plasticity and memory, and continuous destruction of the projecting branches of the cholinergic nuclei in the basal forebrain (84, 85).

### Implications for future clinical research and development of therapeutics

Findings in the current study, if confirmed, could have implications for the conceptualization of Alzheimer pathological cascades, therapeutic targets, and the application of existing and future treatments. The genetic architecture of prognosis in AD, that is fundamental for patients and the design of clinical trials, is less well established than the genetics of susceptibility. *BCHE-K* may join *APOE4* allele frequency, age, and sex as a foundational component of predictive modelling of early AD phenotypes (86, 87). Moreover, the cholinergic hypothesis appears seamlessly interlinked with the amyloid cascade hypothesis. *APOE* and *BCHE* genotypes appear to exert critical influence on the functional activation of glia as indexed by the microglial and astrocyte activation marker, YKL-40, in the CSF. Levels of extracellular cholinergic signaling to cholinoceptive cells, including glia, depend on the enzymatic activity of BuChE, that relies on BuChE levels and polymorphic variation, ApoE levels and polymorphic variation, and soluble Aβ levels. The health of cholinergic neurotransmission and extracellular signaling systems may be crucial to healthy brain aging (88).

The quantitative removal of amyloid pathology with anti-amyloid antibodies is dependent on Fc receptor-mediated phagocytosis and clearance of Aβ (75, 76). Response to this targeted immune activating therapeutic approach may vary depending on the individual’s predominant microglial state. While some beneficial effects might be due to antibody-mediated stimulation of glia with improved performance of homeostatic functions (89), stimulating the clearance of Aβ prior to substantial levels of corticolimbic cholinergic denervation and spreading MTL tau pathology may produce the best outcomes in *APOE4* carriers below the age of 75 years, but this may require intervention in asymptomatic individuals. In addition, longer treatment durations may be necessary in substantial amyloid accumulators, such as *APOE4* homozygotes with *BCHE-K* alleles, to get below the amyloid threshold to prevent or slow further corticolimbic cholinergic denervation and tau pathology.

While retuning of innate immune responses may be required to harness protective and beneficial effects and to attenuate negative effects, the required changes will differ across a genotype, age, and disease stage continuum. Tuning in the wrong direction will simply make matters worse. Considerations may be further complicated by the nuances and complexity of glial cell phenotypes across different brain regions, between adjacent glia, and in different disease contexts (90). The challenge in the development of potential amyloid pathology reducing therapeutics may not lie in simply upregulating the activation state of glia.

## Limitations

Strengths of the study include the well characterized sample of older individuals aged less than 75 years with CSF biomarkers and standardized clinical assessments in expert clinical settings. The limitations of this cross-sectional investigation include its small size and the possibly unrepresentative nature of those enrolled in an interventional clinical trial. Evaluation of the age-at-diagnosis of AD might have benefited from the use of standardized prospective assessments of diagnosis and of onset-age across study sites. Nonetheless, similar genotype group relationships were also demonstrated on the age-at-baseline of study. The findings in the current study appear age-dependent and should not be extrapolated to mild AD patients aged over 75 years. Prospective longitudinal assessment in larger samples is necessary to better evaluate phenotypic evolution along the AD continuum and to confirm and develop these findings.

Although many CSF biomarkers, neuroimaging markers, and clinical assessments were performed, an even more extensive mapping of inflammatory mediators, complement and myelin markers, and CSF ApoE and BChE levels and activity may have been illuminating. The primary biomarker used in this study to index tau pathophysiology (CSF p-tau_181_) may reflect a mix of amyloid and tau pathological changes in the brain (91), and is therefore not a “pure” marker of tau tangle load in the brain. Glia were simplistically ascribed net activated or hypofunctional phenotypes, and as facilitating or limiting amyloid accumulation or tau spread. The association of levels of the astrocyte and microglial activation marker, CSF YKL-40, with transcriptional, morphological, and functional states of glia are not clear, and large multiomic datasets and machine learning may be required to elucidate them.

Although this investigation in study participants of largely European ancestry focused on the influence of specific SNPs in the *APOE* and *BCHE* genes, heterogeneity in the genetic neighborhood of these genes and local *APOE* and *BCHE* haplotypes may be of importance (92). In addition, other than the *K-*variant, the many other genetic variants of *BCHE* were not assessed in the current study (93), and some of those have been shown to unequivocally impact the amyloid cascade (94). Lastly, discerning clinical phenotypes in different genotype and sex subgroups on a variable background of ChE-I therapy and medications with potential anticholinergic properties may be problematic, as such therapy can influence phenotypic expression. For example, *APOE4* carriers— especially those with concomitant *BCHE-K* alleles—are particularly responsive to AChE-I treatment in the mild stage of AD, and the magnitude of attention, processing speed, and amnestic deficits in these individuals may have been partly obscured (32).

## Conclusion

Below the age of 75 years, AD may be more monocausal, without substantial contributions from other age-related pathologies, and the influence of modifying genetic variation on the phenotype of mild AD may be more apparent. In *APOE4* carriers, the presence versus the absence of *BCHE-K* alleles associated with a significantly earlier mean age-at-diagnosis of AD of 6.4 years, a more limbic-amnestic phenotype, and similar accumulations of amyloid and tau pathology but more than 6 years earlier. Thus, in *APOE4* carriers below the age of 75 years, a major contribution to earlier age-at-diagnosis of AD may be concomitant *BCHE-K* alleles. In *APOE4* carriers, *BCHE-K* further reduces the functional activation of glia by increasing cholinergic synaptic signaling from basal forebrain corticolimbic cholinergic neuronal projections and extracellular cholinergic signaling through cell surface α7-nAChRs. The further lowering of glial activation results in earlier amyloid pathology accumulation that, in combination with ApoE4, is particularly damaging to basal forebrain corticolimbic cholinergic neurons. The spread of tau and synaptic pathology from the MTL to other cortical areas parallels the denervation of corticolimbic cholinergic projections, removal of the cholinergic “brake” on cortical glial activation, and the onset and progression of symptoms. The functional activation of glia, the amyloid cascade hypothesis and the cholinergic hypothesis of AD are interwoven. In this mild AD population, the concept has the potential to explain much of the phenotypic heterogeneity, and to enable more appropriate use of existing, emerging, and future therapies. Confirmation of these post hoc findings in larger, prospective, and longitudinal studies is required.

## Data Availability

All data produced in the present work are contained in the manuscript and suppplementary materials (and in addition are available upon reasonable request to the corresponding author)

## Supplementary Material

Raw data (including Mean [SD, SEM], Median [P25, P75], and Min/Max): Table S1. Mild AD phenotype across genotype groups defined by *BCHE-K* allele frequency. Table S2. Mild AD phenotype across genotype groups defined by *APOE4* allele frequency and for *APOE4* homozygotes by the presence of *BCHE-K* alleles. Table S3. Mild AD phenotype across genotype groups defined by *APOE4* and *BCHE-K* carrier status.

Figure S1. Correlations in the overall population and in *APOE4* and *BCHE-K* subgroups of Ng versus YKL-40.

## Ethics committees approving clinical study

## Acknowledgements

We thank the participants and their companions who participated in the study; the sites, and study team from Ionis for executing the study; Michael Moore (Moore Editing, San Diego, CA, USA), who copyedited and styled the manuscript per journal requirements.

## Author contributions

RML was responsible for study design, statistical analysis plan design, and data interpretation, and wrote the initial draft of the manuscript. TD-S critically reviewed, styled, and gave input into the manuscript. CJ oversaw data collection, data analysis, and review of the manuscript. DL and QY advised on statistical analysis plan, performed data analysis, data interpretation, and critical review of the manuscript. KM performed clinical operations and data collection. ALE and DLG designed and conducted biomarker analyses and performed data interpretation and critical review of the manuscript. CJM was the lead investigator and performed data collection, data interpretation, participant recruitment, and critical review of the manuscript. All authors reviewed and provided feedback on the manuscript. The authors had full editorial control of the manuscript and provided their final approval of all content.

## Availability of data and materials

The baseline data that support the findings of this investigation are available in the supplementary materials (Figure S1; Tables S1-S3) and additional data and materials are available upon request.

## Declarations

### Ethics approval and consent to participate

The trial (NCT03186989) was conducted in accordance with Good Clinical Practice Guidelines of the International Council for Harmonisation, and according to the ethical principles outlined in the Declaration of Helsinki.

Patients provided written, informed consent at the time of recruitment. The study was approved by the institutional review board or independent ethics committee at each investigational site; see supplementary materials (Additional File 2).

### Consent for publication

Not applicable.

