## Supplementary table data for "Onset of Alzheimer disease in apolipoprotein ɛ4 carriers is earlier in butyrylcholinesterase K variant carriers"

### Table S1. Mild AD phenotype across genotype groups defined by *BCHE-K allele* frequency.

| Variable | | | 2 *BCHE-K* & 1 *BCHE-K* (n=16) | 2 *BCHE-K* (n=3) | 1 *BCHE-K* (n=13) | No *BCHE-K* (n=29) |
|---|---|---|---|---|---|---|
| **Sex** | Female | | 9 (56.3%) | 2 (66.7%) | 7 (53.8%) | 13 (44.8%) |
| | Male | | 7 (43.8%) | 1 (33.3%) | 6 (46.2%) | 16 (55.2%) |
| **Onset of AD** | Age-at-diagnosis (yrs) | **Mean (SD, SEM)** | **61.63 (6.51, 1.63)** | **59.37 (5.44, 3.14)** | **62.16 (6.82, 1.89)** | **66.02 (5.30, 0.98)** |
| | | Median (P25, P75) | 61.58 (56.64, 66.08) | 57.92 (54.80, 65.39) | 61.89 (56.69, 66.33) | 66.48 (63.10, 70.23) |
| | | (Min, Max) | (49.04, 74.13) | (54.80, 65.39) | (49.04, 74.13) | (47.99, 72.79) |
| | Age-at-baseline (yrs) | **Mean (SD, SEM)** | **62.9 (6.2, 1.6)** | **60.0 (5.6, 3.2)** | **63.6 (6.4, 1.8)** | **67.4 (4.9, 0.9)** |
| | | Median (P25, P75) | 63.0 (59.0, 67.5) | 59.0 (55.0, 66.0) | 63.0 (59.0, 68.0) | 67.0 (66.0, 71.0) |
| | | (Min, Max) | (50.0, 74.0) | (55.0, 66.0) | (50.0, 74.0) | (52.0, 74.0) |
| | Time since AD diagnosis at baseline (yrs) | **Mean (SD, SEM)** | **1.30 (1.18, 0.29)** | **0.63 (0.44, 0.25)** | **1.46 (1.25, 0.35)** | **1.36 (1.38, 0.26)** |
| | | Median (P25, P75) | 1.02 (0.59, 1.88) | 0.61 (0.20, 1.08) | 1.11 (0.67, 2.17) | 1.36 (0.21, 2.37) |
| | | (Min, Max) | (-0.13, 4.37) | (0.20, 1.08) | (-0.13, 4.37) | (-1.04, 4.01) |
| **Neuroimaging** | Hippocampal vol., % of ICV | **Mean (SD, SEM)** | **0.27 (0.04, 0.01)** | **0.27 (0.02, 0.01)** | **0.27 (0.04, 0.01)** | **0.25 (0.04, 0.01)** |
| | | Median (P25, P75) | 0.27 (0.26, 0.29) | 0.27 (0.25, 0.29) | 0.27 (0.27, 0.29) | 0.24 (0.22, 0.27) |
| | | (Min, Max) | (0.17, 0.33) | (0.25, 0.29) | (0.17, 0.33) | (0.19, 0.34) |
| | Ventricular vol., % of ICV | **Mean (SD, SEM)** | **2.72 (1.38, 0.35)** | **1.84 (0.76, 0.44)** | **2.93 (1.43, 0.40)** | **2.82 (1.19, 0.22)** |
| | | Median (P25, P75) | 2.38 (1.81, 3.58) | 1.75 (1.12, 2.65) | 2.80 (2.00, 3.86) | 2.67 (1.94, 3.49) |
| | | (Min, Max) | (1.07, 6.20) | (1.12, 2.65) | (1.07, 6.20) | (1.02, 5.11) |
| **CSF markers** | $A\beta_{40}$, pg/mL | **Mean (SD, SEM)** | **17.44 (3.95, 0.99)** | **18.60 (4.28, 2.47)** | **17.18 (4.01, 1.11)** | **18.20 (4.46, 0.83)** |

| Variable | | 2 *BCHE-K* & 1 *BCHE-K* (n=16) | 2 *BCHE-K* (n=3) | 1 *BCHE-K* (n=13) | No *BCHE-K* (n=29) |
|---|---|---|---|---|---|
| | Median (P25, P75) | 16.76 (14.66, 20.76) | 16.56 (15.73, 23.52) | 16.96 (14.65, 20.52) | 17.68 (16.31, 19.42) |
| | (Min, Max) | (9.95, 23.52) | (15.73, 23.52) | (9.95, 22.74) | (10.73, 28.40) |
| $A\beta_{42}$, pg/mL | **Mean (SD, SEM)** | **676.1 (160.9, 40.2)** | **657.3 (220.2, 127.1)** | **680.5 (155.5, 43.1)** | **713.2 (198.1, 36.8)** |
| | Median (P25, P75) | 653.5 (570.1, 805.0) | 675.8 (428.5, 867.7) | 642.3 (572.0, 804.8) | 687.1 (592.2, 864.6) |
| | (Min, Max) | (428.5, 1039.0) | (428.5, 867.7) | (491.4, 1039.0) | (340.4, 1126.0) |
| $A\beta_{42}$ to $A\beta_{40}$ ratio | **Mean (SD, SEM)** | **0.0400 (0.0075, 0.0019)** | **0.0364 (0.0102, 0.0059)** | **0.0409 (0.0071, 0.0020)** | **0.0389 (0.0075, 0.0014)** |
| | Median (P25, P75) | 0.0423 (0.0342, 0.0462) | 0.0370 (0.0259, 0.0463) | 0.0438 (0.0344, 0.0461) | 0.0385 (0.0335, 0.0446) |
| | (Min, Max) | (0.0259, 0.0504) | (0.0259, 0.0463) | (0.0287, 0.0504) | (0.0255, 0.0517) |
| Total-tau | **Mean (SD, SEM)** | **398.16 (108.96, 27.24)** | **405.18 (139.05, 80.28)** | **396.53 (107.72, 29.88)** | **399.13 (141.17, 26.22)** |
| | Median (P25, P75) | 388.45 (304.88, 480.70) | 330.95 (319.00, 565.60) | 405.60 (290.75, 464.70) | 345.10 (313.45, 454.90) |
| | (Min, Max) | (221.05, 565.60) | (319.00, 565.60) | (221.05, 556.10) | (212.00, 811.75) |
| $p\text{-tau}_{181}$, pg/mL | **Mean (SD, SEM)** | **41.06 (12.91, 3.23)** | **41.95 (13.93, 8.05)** | **40.85 (13.25, 3.68)** | **39.11 (14.30, 2.65)** |
| | Median (P25, P75) | 38.96 (31.52, 51.20) | 36.07 (31.91, 57.86) | 39.36 (31.12, 48.31) | 35.57 (30.29, 44.83) |
| | (Min, Max) | (19.71, 63.98) | (31.91, 57.86) | (19.71, 63.98) | (20.55, 81.28) |
| NfL, pg/mL | **Mean (SD, SEM)** | **1368.151 (416.737, 104.184)** | **1250.411 (270.156, 155.974)** | **1395.322 (447.949, 124.239)** | **1279.087 (395.142, 73.376)** |
| | Median (P25, P75) | 1270.970 (1139.410, 1538.598) | 1213.900 (1000.368, 1536.965) | 1296.260 (1167.610, 1540.230) | 1325.795 (999.923, 1564.960) |
| | (Min, Max) | (630.833, 2391.810) | (1000.368, 1536.965) | (630.833, 2391.810) | (580.969, 2223.485) |
| Ng, pg/mL | **Mean (SD, SEM)** | **559.330 (199.224, 49.806)** | **560.000 (238.627, 137.771)** | **559.176 (200.305, 55.555)** | **508.231 (250.610, 46.537)** |
| | Median (P25, P75) | 541.329 (392.145, 744.592) | 538.994 (332.570, 808.435) | 543.664 (422.006, 738.869) | 470.174 (315.335, 562.083) |
| | (Min, Max) | (237.776, 905.365) | (332.570, 808.435) | (237.776, 905.365) | (194.618, 1306.915) |

| Variable | | | 2 *BCHE-K* & 1 *BCHE-K* (n=16) | 2 *BCHE-K* (n=3) | 1 *BCHE-K* (n=13) | No *BCHE-K* (n=29) |
|---|---|---|---|---|---|---|
| | YKL-40, pg/mL | **Mean (SD, SEM)** | **279704.0 (152934.4, 38233.6)** | **226656.3 (75313.2, 43482.1)** | **291945.8 (165604.8, 45930.5)** | **258422.6 (109116.7, 20262.5)** |
| | | Median (P25, P75) | 230043.5 (147203.5, 382781.8) | 239378.0 (145792.5, 294798.5) | 220709.0 (148614.5, 385381.5) | 236943.0 (202933.5, 300130.5) |
| | | (Min, Max) | (98750.8, 598333.5) | (145792.5, 294798.5) | (98750.8, 598333.5) | (112021.5, 662666.5) |
| Cognition | MMSE Total (0-30) | **Mean (SD, SEM)** | **23.7 (2.4, 0.6)** | **22.3 (1.5, 0.9)** | **24.0 (2.5, 0.7)** | **23.6 (2.2, 0.4)** |
| | | Median (P25, P75) | 24.5 (21.5, 26.0) | 22.0 (21.0, 24.0) | 25.0 (22.0, 26.0) | 23.0 (22.0, 26.0) |
| | | (Min, Max) | (20.0, 27.0) | (21.0, 24.0) | (20.0, 27.0) | (20.0, 27.0) |
| | MMSE Memory (0-6) | **Mean (SD, SEM)** | **4.0 (1.1, 0.3)** | **3.3 (0.6, 0.3)** | **4.2 (1.1, 0.3)** | **4.3 (1.1, 0.2)** |
| | | Median (P25, P75) | 4.0 (3.0, 4.5) | 3.0 (3.0, 4.0) | 4.0 (4.0, 5.0) | 4.0 (3.0, 5.0) |
| | | (Min, Max) | (2.0, 6.0) | (3.0, 4.0) | (2.0, 6.0) | (3.0, 6.0) |
| | MMSE Visual Construction (0-1) | **Mean (SD, SEM)** | **0.8 (0.4, 0.1)** | **0.7 (0.6, 0.3)** | **0.8 (0.4, 0.1)** | **0.6 (0.5, 0.1)** |
| | | Median (P25, P75) | 1.0 (0.5, 1.0) | 1.0 (0.0, 1.0) | 1.0 (1.0, 1.0) | 1.0 (0.0, 1.0) |
| | | (Min, Max) | (0.0, 1.0) | (0.0, 1.0) | (0.0, 1.0) | (0.0, 1.0) |
| | RBANS Total (40-160) | **Mean (SD, SEM)** | **67.5 (11.5, 2.9)** | **63.3 (15.6, 9.0)** | **68.5 (10.9, 3.0)** | **67.1 (12.0, 2.2)** |
| | | Median (P25, P75) | 71.5 (56.0, 77.0) | 61.0 (49.0, 80.0) | 72.0 (57.0, 77.0) | 66.0 (59.0, 76.0) |
| | | (Min, Max) | (49.0, 82.0) | (49.0, 80.0) | (51.0, 82.0) | (49.0, 90.0) |
| | RBANS Delayed Memory (40-154) | **Mean (SD, SEM)** | **57.8 (19.2, 4.8)** | **58.7 (22.7, 13.1)** | **57.6 (19.4, 5.4)** | **51.8 (13.3, 2.5)** |
| | | Median (P25, P75) | 50.0 (44.0, 66.0) | 52.0 (40.0, 84.0) | 48.0 (44.0, 64.0) | 48.0 (44.0, 52.0) |
| | | (Min, Max) | (40.0, 102.0) | (40.0, 84.0) | (40.0, 102.0) | (40.0, 94.0) |
| | RBANS Visuospatial/ Constructional (40-154) | **Mean (SD, SEM)** | **86.9 (24.1, 6.0)** | **79.0 (36.4, 21.0)** | **88.8 (22.1, 6.1)** | **84.2 (20.5, 3.8)** |
| | | Median (P25, P75) | 78.0 | 60.0 | 78.0 | 84.0 |

| Variable | | | 2 *BCHE-K* & 1 *BCHE-K* (n=16) | 2 *BCHE-K* (n=3) | 1 *BCHE-K* (n=13) | No *BCHE-K* (n=29) |
|---|---|---|---|---|---|---|
| | | | (64.0, 105.0) | (56.0, 121.0) | (72.0, 105.0) | (64.0, 102.0) |
| | | (Min, Max) | (56.0, 126.0) | (56.0, 121.0) | (62.0, 126.0) | (50.0, 121.0) |
| | RBANS Immediate Memory (40-154) | **Mean (SD, SEM)** | **62.6 (17.0, 4.2)** | **60.7 (18.0, 10.4)** | **63.1 (17.5, 4.9)** | **68.2 (15.3, 2.8)** |
| | | Median (P25, P75) | 63.0 (49.0, 73.0) | 69.0 (40.0, 73.0) | 57.0 (49.0, 73.0) | 61.0 (57.0, 78.0) |
| | | (Min, Max) | (40.0, 100.0) | (40.0, 73.0) | (40.0, 100.0) | (44.0, 103.0) |
| | RBANS Language (40-154) | **Mean (SD, SEM)** | **88.2 (9.2, 2.3)** | **91.3 (6.4, 3.7)** | **87.5 (9.8, 2.7)** | **87.1 (14.0, 2.6)** |
| | | Median (P25, P75) | 90.0 (82.0, 94.0) | 94.0 (84.0, 96.0) | 90.0 (82.0, 92.0) | 92.0 (83.0, 96.0) |
| | | (Min, Max) | (71.0, 108.0) | (84.0, 96.0) | (71.0, 108.0) | (51.0, 104.0) |
| | RBANS Attention (40-154) | **Mean (SD, SEM)** | **74.4 (14.4, 3.6)** | **58.0 (12.3, 7.1)** | **78.2 (12.3, 3.4)** | **77.7 (15.2, 2.8)** |
| | | Median (P25, P75) | 73.5 (68.0, 85.0) | 53.0 (49.0, 72.0) | 75.0 (72.0, 85.0) | 75.0 (64.0, 85.0) |
| | | (Min, Max) | (49.0, 100.0) | (49.0, 72.0) | (53.0, 100.0) | (53.0, 118.0) |

ICV, intracranial volume; MMSE, Mini-Mental Status Examination; RBANS, Repeatable Battery for the Assessment of Neuropsychological Status; SD, standard deviation; SEM, standard error of the mean.

### Table S2. Mild AD phenotype across genotype groups defined by *APOE4* allele frequency and for *APOE4* homozygotes by the presence of *BCHE-K* alleles.

| Variable | | | 2 *APOE4* & *BCHE-K* (N = 4) | 2 *APOE4* (N = 10) | 1 *APOE4* (N = 23) | No *APOE4* (N = 12) | 1 or 2 *APOE4* (N = 33) |
|---|---|---|---|---|---|---|---|
| **Sex** | Female | | 3 (75.0%) | 6 (60.0%) | 12 (52.2%) | 4 (33.3%) | 18 (54.5%) |
| | Male | | 1 (25.0%) | 4 (40.0%) | 11 (47.8%) | 8 (66.7%) | 15 (45.5%) |
| **Onset of AD** | Age-at-diagnosis (yrs) | **Mean (SD, SEM)** | **60.77 (4.41, 2.20)** | **64.16 (5.38, 1.70)** | **65.21 (5.77, 1.20)** | **63.28 (7.36, 2.12)** | **64.89 (5.59, 0.97)** |
| | | Median (P25, P75) | 61.45 (57.90, 63.64) | 63.03 (61.00, 67.84) | 66.31 (63.10, 69.08) | 62.72 (58.54, 69.31) | 65.83 (62.67, 68.44) |
| | | (Min, Max) | (54.80, 65.39) | (54.80, 72.79) | (49.04, 72.44) | (47.99, 74.13) | (49.04, 72.79) |
| | Age-at-baseline (yrs) | **Mean (SD, SEM)** | **62.0 (4.8, 2.4)** | **65.4 (5.4, 1.7)** | **66.6 (5.6, 1.2)** | **64.6 (6.5, 1.9)** | **66.2 (5.5, 1.0)** |
| | | Median (P25, P75) | 63.5 (59.0, 65.0) | 65.0 (63.0, 68.0) | 67.0 (63.0, 71.0) | 66.0 (59.5, 70.0) | 67.0 (63.0, 70.0) |
| | | (Min, Max) | (55.0, 66.0) | (55.0, 73.0) | (50.0, 74.0) | (52.0, 74.0) | (50.0, 74.0) |
| | Time since AD diagnosis at baseline (yrs) | **Mean (SD, SEM)** | **1.23 (1.24, 0.62)** | **1.24 (1.23, 0.39)** | **1.40 (1.27, 0.26)** | **1.31 (1.49, 0.43)** | **1.35 (1.24, 0.22)** |
| | | Median (P25, P75) | 0.86 (0.41, 2.05) | 0.86 (0.21, 1.65) | 1.36 (0.52, 2.37) | 0.96 (0.21, 2.30) | 1.15 (0.33, 2.37) |
| | | (Min, Max) | (0.20, 3.00) | (0.16, 3.61) | (-1.04, 4.37) | (-0.63, 4.01) | (-1.04, 4.37) |
| **Neuroimaging** | Hippocampal vol., % of ICV | **Mean (SD, SEM)** | **0.26 (0.05, 0.03)** | **0.24 (0.04, 0.01)** | **0.25 (0.04, 0.01)** | **0.28 (0.03, 0.01)** | **0.25 (0.04, 0.01)** |
| | | Median (P25, P75) | 0.28 (0.23, 0.29) | 0.24 (0.23, 0.27) | 0.25 (0.21, 0.27) | 0.29 (0.27, 0.30) | 0.24 (0.22, 0.27) |
| | | (Min, Max) | (0.18, 0.29) | (0.18, 0.29) | (0.17, 0.34) | (0.22, 0.32) | (0.17, 0.34) |
| | Ventricular vol., % of ICV | **Mean (SD, SEM)** | **1.77 (0.73, 0.36)** | **1.84 (0.53, 0.17)** | **3.26 (1.32, 0.28)** | **2.66 (1.08, 0.31)** | 2.83 (1.31, 0.23) |
| | | Median (P25, P75) | 1.66 (1.18, 2.37) | 1.81 (1.36, 2.23) | 2.98 (2.07, 4.34) | 2.49 (1.92, 3.29) | 2.65 (1.94, 3.86) |
| | | (Min, Max) | (1.12, 2.65) | (1.12, 2.65) | (1.02, 6.20) | (1.07, 5.11) | (1.02, 6.20) |
| **CSF markers** | $A\beta_{40}$, pg/mL | **Mean (SD, SEM)** | **16.86** | **17.11** | **17.64** | **19.18** | **17.48** |

| Variable | | 2 *APOE4* & *BCHE-K* (N = 4) | 2 *APOE4* (N = 10) | 1 *APOE4* (N = 23) | No *APOE4* (N = 12) | 1 or 2 *APOE4* (N = 33) |
|---|---|---|---|---|---|---|
| | | **(2.56, 1.28)** | **(1.61, 0.51)** | **(5.02, 1.05)** | **(4.19, 1.21)** | **(4.25, 0.74)** |
| | Median (P25, P75) | 16.14 (15.19, 18.54) | 16.76 (16.54, 18.11) | 16.54 (14.67, 20.99) | 19.43 (16.22, 22.30) | 16.66 (15.73, 19.12) |
| | (Min, Max) | (14.65, 20.52) | (14.65, 20.52) | (9.95, 28.40) | (11.99, 25.34) | (9.95, 28.40) |
| $A\beta_{42}$, pg/mL | **Mean (SD, SEM)** | **553.9 (110.6, 55.3)** | **580.7 (109.6, 34.7)** | **700.2 (188.0, 39.2)** | **799.2 (179.6, 51.9)** | **664.0 (175.4, 30.5)** |
| | Median (P25, P75) | 555.7 (464.5, 643.4) | 587.5 (479.8, 661.5) | 687.1 (582.7, 849.7) | 823.3 (638.0, 900.1) | 661.5 (563.9, 773.6) |
| | (Min, Max) | (428.5, 675.8) | (428.5, 767.3) | (340.4, 1059.5) | (568.3, 1126.0) | (340.4, 1059.5) |
| $A\beta_{42}$ to $A\beta_{40}$ ratio | **Mean (SD, SEM)** | **0.0342 (0.0087, 0.0044)** | **0.0342 (0.0074, 0.0024)** | **0.0399 (0.0063, 0.0013)** | **0.0426 (0.0078, 0.0023)** | **0.0382 (0.0071, 0.0012)** |
| | Median (P25, P75) | 0.0322 (0.0281, 0.0402) | 0.0338 (0.0259, 0.0400) | 0.0409 (0.0347, 0.0446) | 0.0458 (0.0357, 0.0492) | 0.0385 (0.0335, 0.0438) |
| | (Min, Max) | (0.0259, 0.0463) | (0.0255, 0.0463) | (0.0267, 0.0504) | (0.0287, 0.0517) | (0.0255, 0.0504) |
| Total-tau | **Mean (SD, SEM)** | **333.40 (99.21, 49.61)** | **353.51 (95.01, 30.04)** | **414.17 (144.58, 30.15)** | **407.03 (123.68, 35.70)** | **395.79 (133.09, 23.17)** |
| | Median (P25, P75) | 324.98 (270.03, 396.78) | 327.60 (313.45, 345.10) | 384.10 (290.20, 510.25) | 395.68 (308.75, 486.88) | 346.75 (313.45, 464.70) |
| | (Min, Max) | (221.05, 462.60) | (221.05, 566.80) | (212.00, 811.75) | (251.75, 659.30) | (212.00, 811.75) |
| p-$tau_{181}$, pg/mL | **Mean (SD, SEM)** | **34.00 (11.80, 5.90)** | **35.18 (9.31, 2.94)** | **41.05 (15.24, 3.18)** | **41.27 (13.79, 3.98)** | **39.27 (13.84, 2.41)** |
| | Median (P25, P75) | 33.99 (25.81, 42.19) | 32.96 (31.04, 36.07) | 39.36 (28.28, 53.22) | 38.45 (30.01, 51.29) | 36.03 (31.04, 48.31) |
| | (Min, Max) | (19.71, 48.31) | (19.71, 52.59) | (20.55, 81.28) | (23.03, 69.12) | (19.71, 81.28) |
| NfL, pg/mL | **Mean (SD, SEM)** | **1003.178 (264.632, 132.316)** | **1038.510 (319.069, 100.899)** | **1383.129 (438.081, 91.346)** | **1398.905 (296.255, 85.521)** | **1278.699 (431.788, 75.165)** |
| | Median (P25, P75) | 1083.989 (815.600, 1190.755) | 1074.329 (737.715, 1167.610) | 1395.075 (1091.940, 1572.575) | 1358.548 (1178.445, 1615.488) | 1192.395 (1000.368, 1540.230) |
| | (Min, Max) | (630.833, 1213.900) | (630.833, 1718.680) | (580.969, 2391.810) | (999.923, 1992.835) | (580.969, 2391.810) |
| Ng, pg/mL | **Mean (SD, SEM)** | **493.087 (187.517, 93.759)** | **481.615 (119.604, 37.822)** | **546.251 (270.939, 56.495)** | **525.671 (235.097, 67.867)** | **526.664 (235.375, 40.973)** |
| | Median (P25, P75) | 450.454 (347.242, 638.931) | 472.203 (369.620, 538.994) | 479.523 (307.055, 684.491) | 460.332 (338.809, 696.191) | 474.233 (369.620, 582.299) |

| Variable | | | 2 *APOE4* & *BCHE-K* (N = 4) | 2 *APOE4* (N = 10) | 1 *APOE4* (N = 23) | No *APOE4* (N = 12) | 1 or 2 *APOE4* (N = 33) |
|---|---|---|---|---|---|---|---|
| | | (Min, Max) | (332.570, 738.869) | (332.570, 738.869) | (194.618, 1306.915) | (237.776, 1013.229) | (194.618, 1306.915) |
| | YKL-40, pg/mL | **Mean (SD, SEM)** | **190012.7 (86035.8, 43017.9)** | **201544.9 (63834.1, 20186.1)** | **267090.9 (114893.9, 23957.0)** | **317581.8 (161891.7, 46734.1)** | **247228.5 (105627.6, 18387.4)** |
| | | Median (P25, P75) | 183250.8 (122271.6, 257753.8) | 223078.0 (145792.5, 242007.5) | 252314.0 (197204.5, 321401.5) | 256480.3 (204270.8, 433662.3) | 235441.5 (189715.0, 294798.5) |
| | | (Min, Max) | (98750.8, 294798.5) | (98750.8, 294798.5) | (125213.5, 598333.5) | (131158.5, 662666.5) | (98750.8, 598333.5) |
| Cognition | MMSE Total (0-30) | **Mean (SD, SEM)** | **24.3 (2.5, 1.3)** | **24.4 (2.1, 0.7)** | **23.3 (2.3, 0.5)** | **23.7 (2.2, 0.6)** | **23.6 (2.3, 0.4)** |
| | | Median (P25, P75) | 24.5 (22.5, 26.0) | 24.5 (23.0, 26.0) | 23.0 (21.0, 25.0) | 23.5 (21.5, 26.0) | 24.0 (22.0, 26.0) |
| | | (Min, Max) | (21.0, 27.0) | (21.0, 27.0) | (20.0, 27.0) | (21.0, 26.0) | (20.0, 27.0) |
| | MMSE Memory (0-6) | **Mean (SD, SEM)** | **4.0 (1.4, 0.7)** | **4.1 (1.0, 0.3)** | **4.0 (1.1, 0.2)** | **4.6 (1.2, 0.4)** | **4.1 (1.1, 0.2)** |
| | | Median (P25, P75) | 3.5 (3.0, 5.0) | 4.0 (3.0, 5.0) | 4.0 (3.0, 4.0) | 4.5 (3.5, 6.0) | 4.0 (3.0, 4.0) |
| | | (Min, Max) | (3.0, 6.0) | (3.0, 6.0) | (2.0, 6.0) | (3.0, 6.0) | (2.0, 6.0) |
| | MMSE Visual Construction (0-1) | **Mean (SD, SEM)** | **0.8 (0.5, 0.3)** | **0.8 (0.4, 0.1)** | **0.7 (0.5, 0.1)** | **0.5 (0.5, 0.2)** | **0.7 (0.5, 0.1)** |
| | | Median (P25, P75) | 1.0 (0.5, 1.0) | 1.0 (1.0, 1.0) | 1.0 (0.0, 1.0) | 0.5 (0.0, 1.0) | 1.0 (0.0, 1.0) |
| | | (Min, Max) | (0.0, 1.0) | (0.0, 1.0) | (0.0, 1.0) | (0.0, 1.0) | (0.0, 1.0) |
| | RBANS Total (40-160) | **Mean (SD, SEM)** | **72.8 (8.3, 4.2)** | **73.4 (10.5, 3.3)** | **64.0 (11.2, 2.3)** | **68.5 (12.3, 3.5)** | **66.8 (11.7, 2.0)** |
| | | Median (P25, P75) | 75.0 (67.0, 78.5) | 78.0 (61.0, 80.0) | 61.0 (54.0, 73.0) | 67.5 (61.5, 78.5) | 66.0 (57.0, 77.0) |
| | | (Min, Max) | (61.0, 80.0) | (57.0, 85.0) | (49.0, 88.0) | (49.0, 90.0) | (49.0, 88.0) |
| | RBANS Delayed Memory (40-154) | **Mean (SD, SEM)** | **56.0 (19.3, 9.7)** | **52.8 (12.5, 3.9)** | **48.0 (7.4, 1.6)** | **66.3 (22.5, 6.5)** | **49.5 (9.3, 1.6)** |
| | | Median (P25, P75) | 50.0 (44.0, 68.0) | 50.0 (44.0, 56.0) | 48.0 (44.0, 48.0) | 60.0 (48.0, 91.5) | 48.0 (44.0, 52.0) |
| | | (Min, Max) | (40.0, 84.0) | (40.0, 84.0) | (40.0, 68.0) | (40.0, 102.0) | (40.0, 84.0) |
| | RBANS Visuospatial/ Constructional | **Mean (SD, SEM)** | **101.8 (28.8, 14.4)** | **100.8 (20.0, 6.3)** | **81.1 (19.8, 4.1)** | **79.9 (21.7, 6.3)** | **87.1 (21.6, 3.8)** |

| Variable | | 2 *APOE4* & *BCHE-K* (N = 4) | 2 *APOE4* (N = 10) | 1 *APOE4* (N = 23) | No *APOE4* (N = 12) | 1 or 2 *APOE4* (N = 33) |
|---|---|---|---|---|---|---|
| (40-154) | Median (P25, P75) | 113.0 (82.5, 121.0) | 102.5 (84.0, 121.0) | 78.0 (64.0, 105.0) | 79.5 (61.0, 99.0) | 84.0 (69.0, 105.0) |
| | (Min, Max) | (60.0, 121.0) | (60.0, 121.0) | (50.0, 126.0) | (50.0, 116.0) | (50.0, 126.0) |

ICV, intracranial volume; MMSE, Mini-Mental Status Examination; RBANS, Repeatable Battery for the Assessment of Neuropsychological Status; SD, standard deviation; SEM, standard error of the mean.

### Table S3. Mild AD phenotype across genotype groups defined by *APOE4* and *BCHE-K* carrier status.

| Variable | | | *APOE4* & *BCHE-K* (N = 11) | *APOE4* & No-*BCHE-K* (N = 22) | No-*APOE4* & *BCHE-K* (N = 5) | No-*APOE4* & No-*BCHE-K* (N = 7) |
|---|---|---|---|---|---|---|
| **Sex** | Female | | 7 (63.6%) | 11 (50.0%) | 2 (40.0%) | 2 (28.6%) |
| | Male | | 4 (36.4%) | 11 (50.0%) | 3 (60.0%) | 5 (71.4%) |
| **Onset of AD** | Age-at-diagnosis (yrs) | **Mean (SD, SEM)** | **60.61 (6.08, 1.83)** | **67.03 (3.96, 0.84)** | **63.88 (7.58, 3.39)** | **62.84 (7.78, 2.94)** |
| | | Median (P25, P75) | 61.89 (54.80, 65.83) | 67.16 (64.04, 70.63) | 61.28 (57.92, 69.41) | 63.00 (59.16, 69.21) |
| | | (Min, Max) | (49.04, 68.44) | (59.37, 72.79) | (56.69, 74.13) | (47.99, 71.45) |
| | Age-at-baseline (yrs) | **Mean (SD, SEM)** | **62.1 (5.9, 1.8)** | **68.3 (4.0, 0.9)** | **64.8 (7.3, 3.2)** | **64.4 (6.6, 2.5)** |
| | | Median (P25, P75) | 63.0 (59.0, 67.0) | 68.0 (66.0, 72.0) | 62.0 (59.0, 71.0) | 66.0 (60.0, 69.0) |
| | | (Min, Max) | (50.0, 69.0) | (62.0, 74.0) | (58.0, 74.0) | (52.0, 72.0) |
| | Time since AD diagnosis at baseline (yrs) | **Mean (SD, SEM)** | **1.48 (1.34, 0.40)** | **1.28 (1.21, 0.26)** | **0.92 (0.67, 0.30)** | **1.59 (1.89, 0.72)** |
| | | Median (P25, P75) | 0.96 (0.56, 2.40) | 1.45 (0.21, 2.37) | 1.08 (0.72, 1.31) | 0.84 (-0.21, 3.55) |
| | | (Min, Max) | (0.20, 4.37) | (-1.04, 3.61) | (-0.13, 1.59) | (-0.63, 4.01) |
| **Neuroimaging** | Hippocampal vol., % of ICV | **Mean (SD, SEM)** | **0.26 (0.05, 0.01)** | **0.24 (0.03, 0.01)** | **0.28 (0.02, 0.01)** | **0.28 (0.03, 0.01)** |
| | | Median (P25, P75) | 0.27 (0.24, 0.29) | 0.24 (0.21, 0.25) | 0.29 (0.27, 0.30) | 0.28 (0.27, 0.30) |
| | | (Min, Max) | (0.17, 0.33) | (0.19, 0.34) | (0.25, 0.31) | (0.22, 0.32) |
| | Ventricular vol., % of ICV | **Mean (SD, SEM)** | **2.91 (1.54, 0.46)** | **2.79 (1.23, 0.26)** | **2.30 (0.97, 0.44)** | **2.91 (1.15, 0.44)** |
| | | Median (P25, P75) | 2.65 (1.86, 3.86) | 2.59 (1.94, 3.61) | 2.12 (1.75, 3.26) | 2.87 (1.93, 3.49) |
| | | (Min, Max) | (1.12, 6.20) | (1.02, 5.02) | (1.07, 3.31) | (1.92, 5.11) |
| **CSF markers** | $A\beta_{40}$, pg/mL | **Mean (SD, SEM)** | **16.32 (3.20, 0.96)** | **18.06 (4.65, 0.99)** | **19.91 (4.68, 2.09)** | **18.67 (4.11, 1.55)** |

| Variable | | *APOE4* & *BCHE-K* (N = 11) | *APOE4* & No-*BCHE-K* (N = 22) | No-*APOE4* & *BCHE-K* (N = 5) | No-*APOE4* & No-*BCHE-K* (N = 7) |
|---|---|---|---|---|---|
| | Median (P25, P75) | 16.00 (14.65, 19.54) | 16.90 (16.31, 19.12) | 21.87 (19.44, 22.74) | 18.50 (14.76, 21.79) |
| | (Min, Max) | (9.95, 20.99) | (10.73, 28.40) | (11.99, 23.52) | (13.18, 25.34) |
| $A\beta_{42}$, pg/mL | **Mean (SD, SEM)** | **640.5 (138.6, 41.8)** | **675.7 (193.2, 41.2)** | **754.5 (194.7, 87.1)** | **831.2 (176.1, 66.6)** |
| | Median (P25, P75) | 642.3 (500.4, 804.8) | 661.5 (563.9, 773.6) | 669.3 (628.3, 867.7) | 845.8 (647.7, 917.2) |
| | (Min, Max) | (428.5, 849.7) | (340.4, 1059.5) | (568.3, 1039.0) | (598.1, 1126.0) |
| $A\beta_{42}$ to $A\beta_{40}$ ratio | **Mean (SD, SEM)** | **0.0406 (0.0077, 0.0023)** | **0.0369 (0.0066, 0.0014)** | **0.0387 (0.0079, 0.0036)** | **0.0453 (0.0071, 0.0027)** |
| | Median (P25, P75) | 0.0438 (0.0341, 0.0463) | 0.0365 (0.0331, 0.0423) | 0.0370 (0.0344, 0.0461) | 0.0489 (0.0405, 0.0496) |
| | (Min, Max) | (0.0259, 0.0504) | (0.0255, 0.0501) | (0.0287, 0.0475) | (0.0316, 0.0517) |
| Total-tau | **Mean (SD, SEM)** | **389.88 (98.91, 29.82)** | **398.74 (149.34, 31.84)** | **416.37 (139.51, 62.39)** | **400.36 (122.24, 46.20)** |
| | Median (P25, P75) | 371.30 (319.00, 464.70) | 344.03 (309.40, 510.25) | 449.70 (290.75, 524.05) | 392.35 (326.75, 405.05) |
| | (Min, Max) | (221.05, 556.10) | (212.00, 811.75) | (251.75, 565.60) | (290.20, 659.30) |
| p-$tau_{181}$, pg/mL | **Mean (SD, SEM)** | **40.39 (12.21, 3.68)** | **38.71 (14.83, 3.16)** | **42.52 (15.76, 7.05)** | **40.37 (13.46, 5.09)** |
| | Median (P25, P75) | 38.56 (31.91, 48.31) | 33.06 (28.28, 50.94) | 46.53 (29.15, 56.05) | 36.99 (30.87, 41.36) |
| | (Min, Max) | (19.71, 63.98) | (20.55, 81.28) | (23.03, 57.86) | (28.79, 69.12) |
| NfL, pg/mL | **Mean (SD, SEM)** | **1391.727 (498.369, 150.264)** | **1222.185 (394.703, 84.151)** | **1316.283 (159.506, 71.333)** | **1457.920 (366.323, 138.457)** |
| | Median (P25, P75) | 1213.900 (1036.835, 1850.245) | 1177.855 (868.289, 1488.770) | 1296.260 (1245.680, 1391.300) | 1440.645 (1042.665, 1709.570) |
| | (Min, Max) | (630.833, 2391.810) | (580.969, 2223.485) | (1111.210, 1536.965) | (999.923, 1992.835) |
| Ng, pg/mL | **Mean (SD, SEM)** | **567.892 (188.029, 56.693)** | **506.050 (257.391, 54.876)** | **540.495 (244.553, 109.367)** | **515.083 (247.274, 93.461)** |
| | Median (P25, P75) | 538.994 (422.006, 738.869) | 471.601 (307.055, 562.083) | 543.664 (362.283, 750.315) | 440.584 (315.335, 642.066) |
| | (Min, Max) | (332.570, 905.365) | (194.618, 1306.915) | (237.776, 808.435) | (302.237, 1013.229) |

| Variable | | | *APOE4* & *BCHE-K* (N = 11) | *APOE4* & No-*BCHE-K* (N = 22) | No-*APOE4* & *BCHE-K* (N = 5) | No-*APOE4* & No-*BCHE-K* (N = 7) |
|---|---|---|---|---|---|---|
| | YKL-40, pg/mL | **Mean (SD, SEM)** | **247158.4 (143270.3, 43197.6)** | **247263.5 (85011.5, 18124.5)** | **351304.4 (164534.1, 73581.9)** | **293494.2 (168456.1, 63670.4)** |
| | | Median (P25, P75) | 206109.0 (145792.5, 294798.5) | 239475.3 (197204.5, 300613.0) | 380182.0 (239378.0, 487142.5) | 235319.5 (202933.5, 300130.5) |
| | | (Min, Max) | (98750.8, 598333.5) | (112021.5, 487722.0) | (131158.5, 518661.0) | (174219.0, 662666.5) |
| **Cognition** | MMSE Total (0-30) | **Mean (SD, SEM)** | **23.8 (2.5, 0.7)** | **23.5 (2.3, 0.5)** | **23.4 (2.4, 1.1)** | **23.9 (2.3, 0.9)** |
| | | Median (P25, P75) | 25.0 (21.0, 26.0) | 23.0 (22.0, 26.0) | 22.0 (22.0, 26.0) | 24.0 (21.0, 26.0) |
| | | (Min, Max) | (20.0, 27.0) | (20.0, 27.0) | (21.0, 26.0) | (21.0, 26.0) |
| | MMSE Memory (0-6) | **Mean (SD, SEM)** | **3.9 (1.0, 0.3)** | **4.1 (1.1, 0.2)** | **4.2 (1.3, 0.6)** | **4.9 (1.2, 0.5)** |
| | | Median (P25, P75) | 4.0 (3.0, 4.0) | 4.0 (3.0, 5.0) | 4.0 (3.0, 5.0) | 5.0 (4.0, 6.0) |
| | | (Min, Max) | (2.0, 6.0) | (3.0, 6.0) | (3.0, 6.0) | (3.0, 6.0) |
| | MMSE Visual Construction (0-1) | **Mean (SD, SEM)** | **0.7 (0.5, 0.1)** | **0.7 (0.5, 0.1)** | **0.8 (0.4, 0.2)** | **0.3 (0.5, 0.2)** |
| | | Median (P25, P75) | 1.0 (0.0, 1.0) | 1.0 (0.0, 1.0) | 1.0 (1.0, 1.0) | 0.0 (0.0, 1.0) |
| | | (Min, Max) | (0.0, 1.0) | (0.0, 1.0) | (0.0, 1.0) | (0.0, 1.0) |
| | RBANS Total (40-160) | **Mean (SD, SEM)** | **65.5 (10.5, 3.2)** | **67.5 (12.4, 2.6)** | **72.0 (13.4, 6.0)** | **66.0 (11.8, 4.5)** |
| | | Median (P25, P75) | 64.0 (55.0, 77.0) | 68.5 (57.0, 79.0) | 77.0 (72.0, 80.0) | 62.0 (61.0, 68.0) |
| | | (Min, Max) | (51.0, 80.0) | (49.0, 88.0) | (49.0, 82.0) | (52.0, 90.0) |
| | RBANS Delayed Memory (40-154) | **Mean (SD, SEM)** | **52.7 (13.7, 4.1)** | **47.8 (5.9, 1.2)** | **69.0 (26.2, 11.7)** | **64.3 (21.5, 8.1)** |
| | | Median (P25, P75) | 48.0 (44.0, 64.0) | 48.0 (44.0, 52.0) | 56.0 (56.0, 91.0) | 64.0 (48.0, 92.0) |
| | | (Min, Max) | (40.0, 84.0) | (40.0, 60.0) | (40.0, 102.0) | (40.0, 94.0) |
| | RBANS Visuospatial/ Constructional (40-154) | **Mean (SD, SEM)** | **90.4 (25.6, 7.7)** | **85.4 (19.8, 4.2)** | **79.4 (21.1, 9.4)** | **80.3 (23.9, 9.0)** |
| | | Median (P25, P75) | 78.0 | 84.0 | 78.0 | 81.0 |

| Variable | | *APOE4* & *BCHE-K* (N = 11) | *APOE4* & No-*BCHE-K* (N = 22) | No-*APOE4* & *BCHE-K* (N = 5) | No-*APOE4* & No-*BCHE-K* (N = 7) |
|---|---|---|---|---|---|
| | | (64.0, 121.0) | (69.0, 105.0) | (62.0, 96.0) | (60.0, 102.0) |
| | (Min, Max) | (60.0, 126.0) | (50.0, 121.0) | (56.0, 105.0) | (50.0, 116.0) |
| RBANS Immediate Memory (40-154) | **Mean (SD, SEM)** | **58.6 (12.3, 3.7)** | **69.5 (16.3, 3.5)** | **71.4 (23.8, 10.6)** | **64.0 (11.9, 4.5)** |
| | Median (P25, P75) | 57.0 (49.0, 69.0) | 67.0 (57.0, 81.0) | 73.0 (57.0, 87.0) | 57.0 (53.0, 78.0) |
| | (Min, Max) | (40.0, 73.0) | (44.0, 103.0) | (40.0, 100.0) | (53.0, 81.0) |
| RBANS Language (40-154) | **Mean (SD, SEM)** | **88.5 (10.6, 3.2)** | **88.1 (13.3, 2.8)** | **87.6 (6.0, 2.7)** | **84.0 (16.7, 6.3)** |
| | Median (P25, P75) | 90.0 (82.0, 94.0) | 92.0 (83.0, 96.0) | 85.0 (84.0, 90.0) | 87.0 (64.0, 99.0) |
| | (Min, Max) | (71.0, 108.0) | (51.0, 104.0) | (82.0, 97.0) | (60.0, 104.0) |
| RBANS Attention (40-154) | **Mean (SD, SEM)** | **72.6 (14.3, 4.3)** | **79.5 (16.6, 3.5)** | **78.4 (15.4, 6.9)** | **71.9 (8.1, 3.1)** |
| | Median (P25, P75) | 72.0 (68.0, 85.0) | 79.0 (64.0, 91.0) | 85.0 (75.0, 88.0) | 72.0 (64.0, 75.0) |
| | (Min, Max) | (49.0, 100.0) | (53.0, 118.0) | (53.0, 91.0) | (60.0, 85.0) |

ICV, intracranial volume; MMSE, Mini-Mental Status Examination; RBANS, Repeatable Battery for the Assessment of Neuropsychological Status; SD, standard deviation; SEM, standard error of the mean.

**Figure S1.** Correlations in the overall population and in *APOE4* and *BCHE-K* subgroups of Ng versus YKL-40.

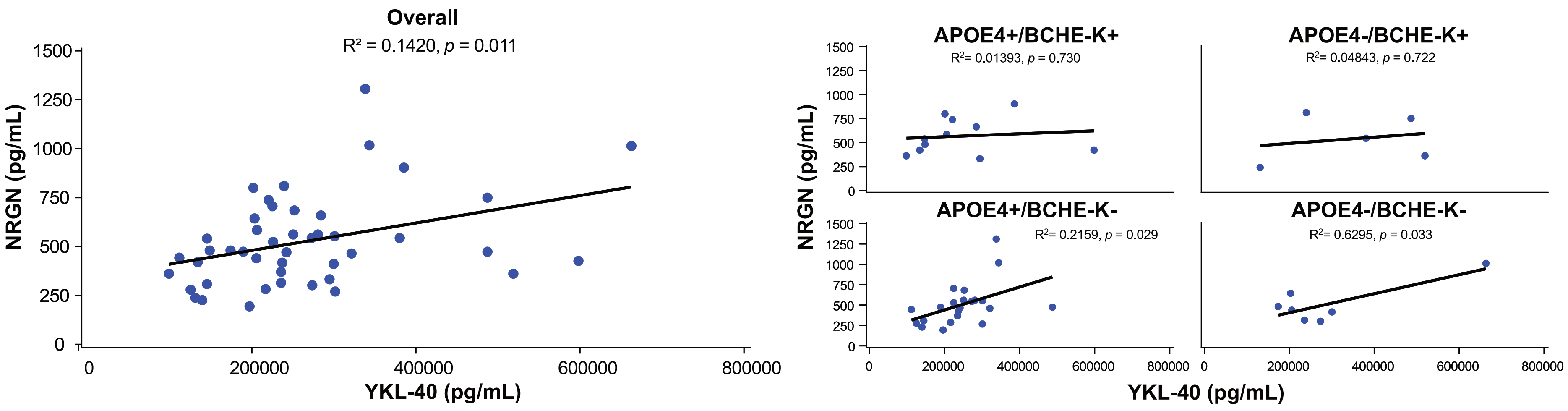

### Ethics committees approving clinical study

| Principal Investigator | IRB/Ethic Committee | Reference number |
|---|---|---|
| Dr Albert Ludolph | Ethikkommission der Universität Ulm | 412/16 |
| Dr Siegfried Muhlack | Ethik-Kommission der Ruhr-Universität Bochum | 16-5929 |
| Dr Catherine Mummery | London-Central Research Ethics Committee Manchester HRA Centre | 17/LO/0440 |
| Dr Simon Ducharme | MUHC Neurosciences Research Ethics Board | 2017-3206 |
| Dr Juha Rinne | National Committee on Medical Research Ethics | 73/06.00.01/2017 |
| Dr Ralf Bodenschatz | Ethikkommission der Sächsischen Landesärztekammer | EK-AMG-MCB-155/16-1 |
| Dr Peter Paul de Deyn | Central Committee on Research Involving Human Subjects | NL60032.000.16 |
| Dr Anne Borjesson Hansen | Regionala etikprövningsnämnden i Stockholm Karolinska Institutet i Solna | 2017/300-31 |
| Dr Michael Jonsson | Regionala etikprövningsnämnden i Stockholm Karolinska Institutet i Solna | 2017/300-31 |
| Dr Daniel Blackburn | London-Central Research Ethics Committee Manchester HRA Centre | 17/LO/0440 |
| Dr Anja Schneider | Ethikkommission an der Medizinischen Fakultät der Rheinischen Friedrich-Wilhelms-Universität Bonn | 035/18-AMG |
| Dr Phillipus Scheltens | Central Committee on Research Involving Human Subjects | NL60032.000.16 |
